# Maternal depression and child human capital: A genetic instrumental-variable approach

**DOI:** 10.1101/2021.02.11.21251547

**Authors:** Giorgia Menta, Anthony Lepinteur, Andrew E. Clark, Simone Ghislandi, Conchita D’Ambrosio

**Affiliations:** University of Luxembourg; Paris School of Economics - CNRS; Bocconi University

**Keywords:** Mendelian Randomisation, Maternal Depression, Human Capital, Instrumental Variables, ALSPAC

## Abstract

We here address the causal relationship between maternal depression and child human capital using UK cohort data. We exploit the conditionally-exogenous variation in mothers’ genomes in an instrumental-variable approach, and describe the conditions under which mother’s genetic variants can be used as valid instruments. An additional episode of maternal depression between the child’s birth up to age nine reduces both their cognitive and non-cognitive skills by 20 to 45% of a SD throughout adolescence. Our results are robust to a battery of sensitivity tests addressing, among others, concerns about pleiotropy and the maternal transmission of genes to her child.

## 1. Introduction

The prevalence of mental-health disorders has been rising steadily for over two decades (Stansfeld *et al*., 2016), and these are now estimated to affect over 20% of the population in the UK (www.mind.org.uk) and the US (www.nami.org/mhstats). Depression is one of the most common of these disorders. A vast literature has documented worse outcomes for the depressed in terms of not only health, but also employment and earnings (Zimmerman and Katon, 2005; Fletcher, 2013; Banerjee *et al*., 2017; Hakulinen *et al*., 2019), productivity (Bubonya *et al*., 2017), marital status and marital satisfaction (Gotlib *et al*., 1998), and parenting style (Kiernan and Huerta, 2008). Major Depressive Disorder has been identified as the largest worldwide contributor to years lost to disability (Prince *et al*., 2007).

Depression in addition, likely also spills over onto others. There is a great deal of work on the intergenerational correlation between parental and child depression (see Gotlib *et al*., 2020, for a recent summary). We here consider the consequences of maternal depression on child human-capital in unique British birth-cohort data, beyond the intergenerational inheritance of the genes associated with depression.

While a broad range of descriptive evidence has underlined the negative association between maternal depression and child outcomes (see Goodman *et al*., 2011, and O’Hara and McCabe, 2013, for meta-analyses and reviews of the psychological literature), there has been little causal analysis of this intergenerational link. One exception is Dahlen (2016), who uses non-parametric bounds to estimate ranges of the negative causal impact of maternal depression on the test scores and socioemotional outcomes of US kindergarten children. More notably, von Hinke *et al*. (2019) rely on unexpected life experiences (the illness or death of friends and family members) to isolate the effect of perinatal maternal depression on children’s cognitive and non-cognitive skills in a UK birth cohort (ALSPAC; the same dataset that we use here). They find that mother’s worse mental health around birth negatively affects their children’s non-cognitive skills, with the effects fading away as the child approaches adolescence. No effect is found on cognitive outcomes.

A small number of contributions have focused on the beneficial causal effects of the successful treatment of depressed mothers. Perry (2008), exploiting the arguably-exogenous variation in US primary-care physicians’ propensity to diagnose depression, shows that treating maternal depression improved children’s asthma outcomes. Using data from a randomised controlled trial, Baranov *et al*. (2020) find that prenatally-depressed mothers in rural Pakistan who were offered psychotherapy had better mental-health outcomes, and invested more time and money in their children (although there is only limited evidence that this investment improved child-development outcomes).

The causal link between parental mental health and child outcomes is of primary policy importance, but is in general not particularly easy to establish. The interplay between maternal mental health and child human-capital development is complex and subject to potential endogeneity concerns. For instance, poor child school performance or behavioural problems might themselves produce maternal depression; alternatively, environmental variables (shared by parents and children who live in the same household), such as local public goods or criminality, could feed through to both parental mental health and child outcomes. In both cases it is difficult to establish causality.

We here address endogeneity via recent advances in Epidemiology and Molecular Genetics. In particular, we adopt a genetic instrumental-variable approach (similar to DiPrete *et al*., 2018), and instrument maternal depression using a synthetic measure (the polygenic score) based on the mother’s genetic variants that are robustly associated with the trait of depression.

Our empirical analysis is based on genetic and socio-economic information on mother-child pairs from the Avon Longitudinal Study of Parents and Children (ALSPAC), a UK-based cohort study that recruited about 14,000 pregnant mothers in the early 1990s. The key explanatory variable is reported maternal depression: this is a summary measure from the answers mothers give to questions about recent depression in seven different data waves from childbirth up to child age nine. We instrument this cumulative depression score by the polygenic score (PGS) for maternal depression, using Genome-Wide Association Studies (GWAS) summary statistics from the depression meta-analysis in Turley *et al*. (2018). Our methodological approach is similar to that in von Hinke *et al*. (2016), who illustrate the assumptions under which an individual’s genetic variants can be used as instrumental variables for that individual’s traits (in their empirical application, child fat mass). Our question differs from theirs, as the trait we instrument (depression) and the outcome (human capital) refer to different individuals (respectively, the mother and her child). In this intergenerational analysis, additional concerns need to be addressed, such as those deriving from genetic inheritance that we will discuss below.

Following the human-capital development and skill-formation literature (see, for example, Cunha and Heckman, 2008), we consider child cognitive and non-cognitive skills as human capital components. The cognitive element is given by the measurement of child skills and knowledge at different stages of compulsory education in the UK. We in particular analyse the child’s average Key Stage test-scores at ages 11 and 14, and their total GCSE score at age 16 (at the end of compulsory education); all three of these test scores come from administrative data. Non-cognitive skills come from the child’s score from the questions in the Strengths and Difficulties Questionnaire (as reported by their principal carer) at child ages 11, 13 and 16.

The genetic instrument allows us to isolate an exogenous change in maternal depression up to child age nine and establish its causal impact on the child’s later human capital. We find that one additional episode of maternal depression (out of the seven recorded) has a persistent negative impact on both cognitive and non-cognitive skills, with an effect size of around 20% of a standard deviation for the former and 40% for the latter.

Our identification strategy relies on a number of assumptions: while the relevance of the mother’s PGS in predicting her depression can be formally tested, the genetic nature of this instrument calls for a more thorough investigation of the exclusion restriction. We illustrate the potential pathways that may compromise identification here, and discuss some ways in which these concerns can be addressed. Pleiotropy (when one genetic variant can explain a number of different traits) is arguably the main issue with genetic instrumentation in general. The intergenerational nature of our research introduces a second potential problem, that of genetic inheritance: as the child inherits about 50% of each parent’s genetic variants, the direct effect of the child’s inherited genetic variants may confound the relationship between mother’s instrumented depression and child human capital. Child outcomes will be affected by the child’s own depression, and this is partly due to the genetic propensity for depression that was inherited from the mother. But this is not what we understand by asking if depressed mothers affect their children’s outcomes: we here rather wish to establish the effect of maternal depression net of genetic inheritance.

We tackle some of the pleiotropic concerns by controlling for a set of maternal and child traits that might be affected by the genetic variants used in the construction of the PGS, and that are in turn likely to affect human-capital development (e.g. educational attainment and fertility decisions). Following Lawlor *et al*. (2017) and DiPrete *et al*. (2018), we control for genetic inheritance by holding constant the child’s own PGS for depression, as well as their PGSs for cognitive and non-cognitive skills. Our results are robust to these and other sensitivity tests.

The remainder of this paper is organised as follows. Section 2 describes the birth-cohort data that we use. Section 3 then provides an overview of the conditions under which genetic variants can be used as instrumental variables in observational data, and considers the specific issues when the treatment and the outcome refer to different individuals who are genetically-related. The main results, of a sizeable causal effect of maternal depression in childhood on adolescent children’s cognitive and non-cognitive skills, and a variety of robustness checks, appear in Section 4. Last, Section 5 concludes.

## 2. Data: The Avon Longitudinal Study of Parents and Children

We will use mother’s genetic information as an instrument to establish the causal effect of her depression on her children’s cognitive and non-cognitive outcomes. The data requirements to carry out this analysis are stringent. We need information on mother’s reported depression during her child’s young years, the adolescent outcomes of her child, and both the the mother’s and the child’s genotype. Few datasets contain all of this information. One that does is the Avon Longitudinal Study of Parents and Children (ALSPAC) survey, also known as ‘The Children of the 90s’.

ALSPAC is an English birth-cohort study designed to investigate the influence of environmental, genetic, and socio-economic variables on health and development over the life course. Over 14,000 pregnant women who were due to give birth between April 1991 and December 1992 in the county of Avon (Bristol and its surrounding areas) were recruited. These women and their families have been followed ever since, even if they move out of the original recruitment area (see www.bristol.ac.uk/alspac/). The pregnancy outcomes of the participants resulted in a total of 14,062 live births, with 13,988 children surviving their first year. The sample is broadly representative of the early 1990s UK population of mothers with children under age one, although higher socio-economic status groups as well as Whites are over-represented (see Fraser *et al*., 2013, and Boyd *et al*., 2013, for a full description of the cohort profile). The study includes detailed information about the family environment, as well as indicators of child development, wellbeing and skills over time, and rich information on the parents’ characteristics and background.^1^ Biological samples from the children and their parents were collected at different points in time, allowing for DNA genotyping. We here use imputed genotype data from around 9,000 children and their mothers (Taylor *et al*., 2018, provide technical details on the genotyping technology, imputation, and quality control in ALSPAC).

When the child was aged 8 months and 2, 3, 4, 5, 6 and 9 years, their mothers were asked whether they had experienced depression since the last interview in which they were asked about their health (or since the birth of the child the first time this question was asked). Although the wording of the question changed slightly across waves, the potential responses were the same: “Yes and consulted a doctor”, “Yes but did not consult a doctor” and “No”. We consider a mother to have had an episode of depression between two periods if she replied “Yes and consulted a doctor” or “Yes but did not consult a doctor”. We combine these seven reported depression scores to produce an index of reported maternal depression from the child’s birth to the child’s ninth birthday, with index values running from zero to seven.

Our child non-cognitive skill measures come from the Strengths and Difficulties Questionnaire (SDQ) (as used in Flèche, 2017; Briole *et al*., 2020; and Clark *et al*., 2021). The SDQ is a 25-question behavioural-screening tool for children, including questions on whether the child is considerate of others, and her concentration span, worries and fears, degree of obedience, and social isolation (Goodman, 1997). The full list of the SDQ items appears in Appendix Table A1. The main carer (this is the mother in the vast majority of cases) was asked to rate the child’s SDQ seven times between child ages 4 and 16. We will relate maternal depression during the child’s first 9 years to the child’s subsequent SDQ scores at ages 11, 13 and 16.

The 25 SDQ items are split up into five sub-scales covering emotional problems, peer problems, conduct problems, hyperactivity/inattention and pro-social behaviour. Consistent with Goodman *et al*. (2010) and the SDQ scores produced by ALSPAC, our main analysis will use the total SDQ score, which is the sum of the first four sub-scales. We code total SDQ so that higher values represent better outcomes (i.e. strengths rather than difficulties). In the robustness checks (Section 4.4.2), we will consider additional non-cognitive skill measures to test for convergent validity (teacher-reported SDQ scores, and an alternative measure of non-cognitive skills from the Short Moods and Feelings Questionnaire, SMFQ, reported by the main carer).

Child cognitive development is measured by their national exam results in linked administrative data from the UK National Pupil Database. We use the average Key Stage fine-grading test-scores at ages 11 and 14 and the total GCSE score in all of the exams that the child took at the end of compulsory education at age 16.^2^

The next section first sets out the principle of using genotype data as an instrument, and then describes the way in which we will apply this method using ALSPAC data.

## 3. A Genetic Instrumental-Variables Approach

### 3.1. Mendelian Randomisation

Establishing causality in non-experimental data is very often challenging, and particularly so for variables that are unlikely to be targeted by policies or be subject to quasi-experimental variation. One recent approach in Social Sciences and Epidemiology is Mendelian Randomisation (MR). This term refers to Mendel’s Laws of Segregation and Independent Assortment, which are involved in the formation of reproductive cells (i.e. gametes) through meiosis and which ensure genetic variability across individuals. Traits that are regulated by one gene are defined by a sequence of two alleles (one inherited from each parent); the Law of Segregation states that each individual has a 50% chance of inheriting one of the two maternal (paternal) alleles for a given gene. The Law of Independent Assortment, on the other hand, ensures that alleles for different traits are passed on independently of each other.^3^ As a result, conditional on the parental genotypes, the child’s genotype can be seen as the outcome of a lottery.^4^

MR in practice refers to a variety of different approaches, the common denominator being the use of genetic variants as instrumental variables for a given endogenous trait (see Koellinger and De Vlaming, 2019, and Hemani *et al*., 2018, for reviews of some recent developments). While some traits can be linked to a clear small set of genetic variants through well-characterised biological pathways (this is the case for severe health problems, such as Huntington’s disease), most traits that interest economists and other social scientists (e.g. socio-economic status, education, and subjective well-being) are highly polygenic and, as such, involve a greater degree of genetic complexity. The burgeoning literature on large-scale GWAS, which aims to estimate the relationship between a given trait and known genetic variants (typically Single-Nucleotide Polymorphisms, or SNPs) in large samples, has brought about significant advances in the understanding of the genetic architecture of genetically-complex traits such as education (Lee *et al*., 2018; Demange *et al*., 2021), depression (Okbay *et al*., 2016; Turley *et al*., 2018) and risk behaviour (Karlsson Linnér *et al*., 2019).

One issue with the use of genetic variants of complex traits as instrumental variables is weak instruments, as each single SNP identified in a GWAS likely has only relatively little predictive power on its own. Polygenic scores have then come into widespread use as linear combinations of all of the relevant genetic markers into one synthetic measure (Appendix B provides more details on the PGS and its functional form), capturing a greater portion of trait variance as compared to single SNPs (DiPrete *et al*., 2018; Davies *et al*., 2015).

We now consider the various relationships between maternal genes and her child’s outcomes, and how these can be addressed to establish a plausible causal relationship.

### 3.2. Instrumental Variable Assumptions in the Context of Genetic Instruments

While others have laid down the assumptions for drawing inference from genetic instruments within the same individual (notably von Hinke *et al*., 2016), we here consider instrumentation between parent and child, as illustrated by the solid black lines in Figure 1. We aim to measure the causal effect of a mother’s trait *D*_*M*_ on her child’s outcome *Y*_*C*_ (i.e. the value of the parameter *β*), where 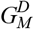 is a vector of independent genetic variants of the mother that are robustly associated with this trait *D*_*M*_. In the ALSPAC analysis that we undertake here, *D*_*M*_ is maternal depression between child birth and child age nine, *Y*_*C*_ the adolescent-child’s human capital, and 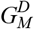 the mother’s PGS for depression, based on the 88 most-relevant SNPs (p-value threshold of 10^−6^) derived from the single-trait meta-analysis in Turley *et al*. (2018). The results throughout the paper are robust to the use of a more-stringent threshold, identifying what are called genome-wide significant SNPs, with a p-value threshold of 5×10^−8^.

**Figure 1:**
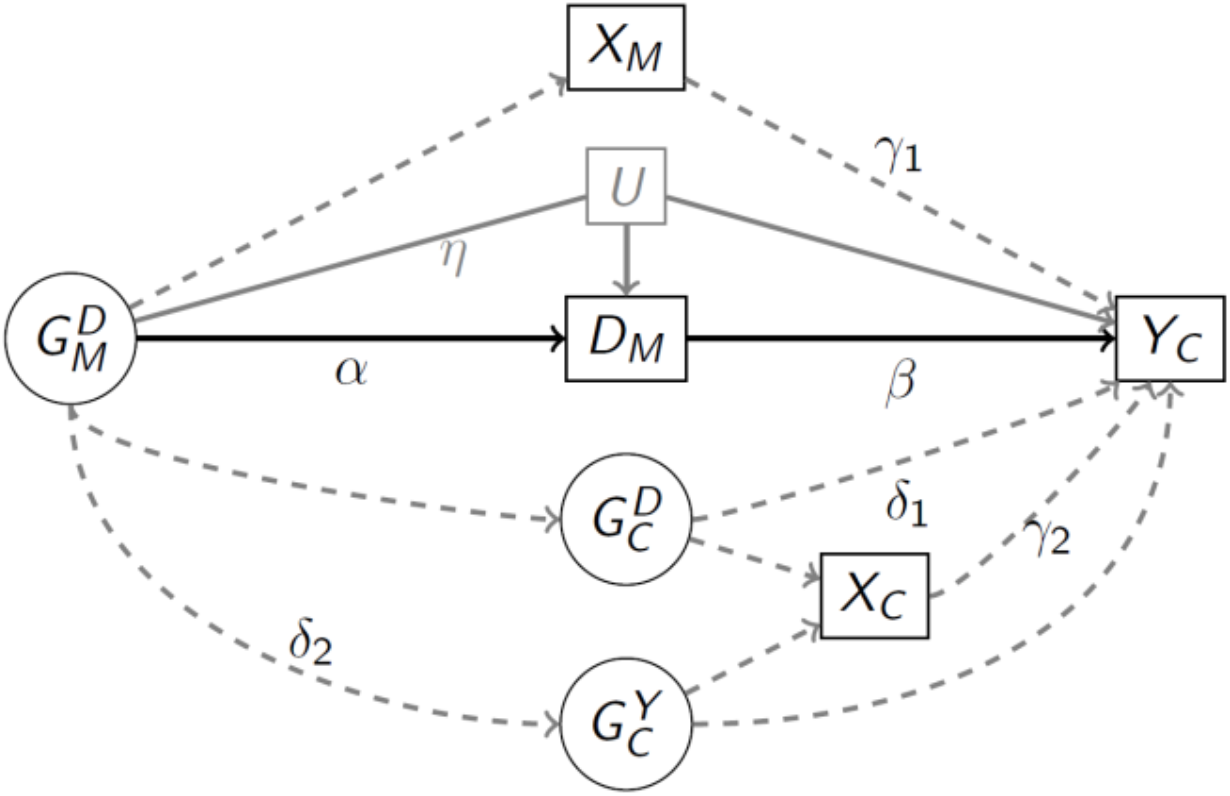
A Directed Acyclic Graph Illustrating the IV Setup and its Assumptions *Notes*: The solid black lines depict the standard IV setup, where 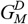 is a (vector of) instrument(s) for a maternal trait *D*_*M*_ and *Y*_*C*_ is the child-level outcome of interest. *U* is a set of unobservable confounders of the trait-outcome association that should not be correlated with 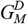 (the independence assumption, i.e. *η* = 0). *X*_*M*_ is a set of maternal traits that are influenced by 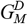 through horizontal pleiotropy or other confounding pathways (e.g. genetic nurture) and have an impact on *Y*_*C*_, thus violating the exclusion restriction. The identification issues in the bottom half of the figure reflect genetic inheritance (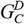 and 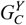 are, respectively, the child’s genetic variants for traits *D* and *Y*). Lines with arrows at the end represent causal relationships; the line between 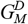 and *U* does not have an arrow and therefore reflects a correlational relationship.

Just as in a standard instrumental variables (IV) analysis, the validity of the identification strategy relies on the following assumptions:

– *Relevance*: the genetic variants 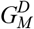 are correlated with the trait *D*_*M*_ (*α* ≠ *0*).
– *Independence*: the 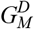 are not correlated with any confounders (*U*) of the association between the mother’s trait and the child outcome (*η* = 0).
– *Exclusion restriction*: the 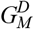 are causally related to the outcome *Y*_*C*_ only through the trait *D*_*M*_ (so that *β* is not confounded by any of the dashed grey lines in Figure 1).

The relevance assumption, while being the easiest to prove in most IV contexts, is particularly straightforward in the case of genetic instrumental variables. The task of identifying which genetic variants are robustly associated with a given trait is typically left to summary data from published GWAS (see Appendix B for further details). As noted above, single genetic variants *per se* might not be sufficiently strong predictors of a trait, especially when the latter is genetically-complex. In these cases, it is more appropriate to use synthetic measures such as the PGS to avoid weak-instrument problems.

The independence assumption is typically assumed to hold in the context of MR due to the randomness of genetic variants, with very few exceptions suggesting otherwise (e.g. Koellinger and De Vlaming, 2019). It is worth underlining, however, that the mother’s genotype can be considered as truly random only when conditioning on the maternal grandparents’ genotype. In practice, for data-availability reasons, it is seldom possible to partial out the genes of the mother’s parents when analysing 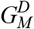. Some common established good practices in MR analyses are controlling for population stratification^5^ and documenting the absence of systematic correlations between the instrument and observable confounders (Smith *et al*., 2007; Boef *et al*., 2015).

In the context of multivariate regressions, controlling for a selected set of grandparental traits, as well as environmental characteristics, should also attenuate the concerns regarding the independence assumption. Consider, as an illustration of *U* in Figure 1, the potential influence of grandparental depression. Depressed grandparents are first more likely to have genetic variants associated with depression: via genetic inheritance, their daughters will then also likely display a higher PGS for depression (in Figure 1 this would translate into *η* ≠ 0). In addition, grandparental depression may increase the chances of their daughter’s depression through non-genetic pathways, e.g. by increasing familial stress and anxiety (this is represented by the line from *U* to *D*_*M*_ in Figure 1). Last, grandparental depression can affect child outcomes directly, as depicted in the line from *U* to *Y*_*C*_: this could reflect, for example, the crowding-out effect of the time that mothers with depressed parents can dedicate to their children. As they may simultaneously affect all of the variables of interest (via the three unbroken grey lines in Figure 1), not controlling for grandparental genes and/or their associated traits can violate the independence assumption.^6^ Introducing controls for the depression of both of the grandparents, as well as for other grandparental traits and environmental characteristics, can attenuate the bias in this case.

The exclusion restriction is well-known to be the most problematic assumption in all IV setups, and this is particularly true in the context of MR (Koellinger and De Vlaming, 2019).

In our mother-child framework, phenomena such as horizontal pleiotropy and genetic inheritance can link 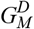 and *Y*_*C*_ through pathways other than *D*_*M*_.

Under horizontal pleiotropy, an individual’s genetic variant directly affects two or more of her traits through separate biological pathways (e.g. the genetic variants causing maternal depression might also affect other maternal traits, such as educational attainment). This will pose identification problems if these additional traits affected by 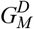 (the *X*_*M*_ in Figure 1) are correlated with the outcome of interest (*γ*_*1*_ ≠ *0*).

One simple way to account for the confounding effects of the *X*_*M*_ in Figure 1 is to control for them. While this may sound rather trivial, most MR applications are actually bivariate associations (accompanied, in most cases, by statistical tools to account for pleiotropy), typically due to data limitations. We do of course need to be careful when controlling for maternal traits: while some might indeed capture part of the observable pleiotropic effects, they can also partly mediate the relationship between *D*_*M*_ and *Y*_*C*_ and, as such, be ‘bad controls’ (Angrist and Pischke, 2008). In relation to Figure 1, holding ‘bad controls’ constant would lead to the attenuation of the estimated value of *β*.

Genetic inheritance also poses a problem for identification. Each child inherits 50% of each parent’s genetic variants. This produces the path from 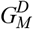 to 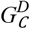, the child’s genetic variants that are associated with child trait *D*, in Figure 1. There are then two pathways from 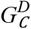 to the child’s outcome. The first is the direct biological pathway from 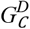 to *Y*_*C*_ (*δ*_1_ ≠ *0*); the second is due to vertical pleiotropy (*γ*_2_ ≠ *0*), i.e. the effect of 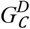 on *Y*_*C*_ that is mediated by one or more child traits (*X*_*C*_).

The issues around genetic inheritance might not only concern the transmission of the genetic variants for depression. The child’s genetic variants explaining *Y*_*C*_, 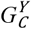 may also partly be inherited from the mother’s 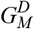 and/or result from linkage disequilibrium (LD from here onwards; see footnote 3 for the definition) with it (*δ*_*2*_ ≠ *0*). This is important here, as mental health and cognitive achievement partly share the same genetic aetiology (see Rajagopal *et al*., 2020). Similarly to 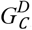, the vector 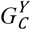 can affect the child outcomes either directly or via vertical pleiotropy (*γ*_*2*_ ≠ *0*).

Were the genetic data detailed enough, we could deal with all the concerns arising from genetic inheritance by using only the mother’s non-transmitted alleles as instruments: mechanically, there would then be no correlation between the mother’s and the child’s genotypes (unless there is assortative matching between the parents over trait *D* and/or *Y*). Another possibility, which is what we do here, is to control for the child’s genotypes, so as to hold constant all of the pathways between 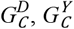 and *Y*_*C*._ We will in addition control for the child’s traits, *X*_*C*_, in case these partly result from genetic variants other than 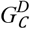 and 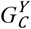. We are to the best of our knowledge the first to be able to control for both the child’s PGS for depression, 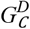 (Lawlor *et al*., 2017), and cognitive (non-cognitive) skills, 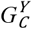 (DiPrete *et al*., 2018),^7^ in the empirical analysis. Note that once we have controlled for the relevant child PGSs, the residual part of the bivariate pathway between *X*_*M*_ and *Y*_*C*_ in Figure 1, *γ*_*1*_, reflects genetic nurture (Kong *et al*., 2018), i.e. the effect of the maternal traits caused by the part of 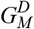 that is not inherited by the child.

We next describe the equations that to be estimated using ALSPAC data.

### 3.3. Empirical Strategy

We address endogeneity by estimating the following Two-Stage Least Squares (2SLS) regressions using a genetic instrument:

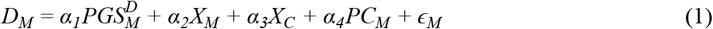

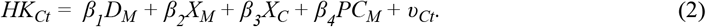

In Equation (1), *D*_*M*_ is the number of self-reported episodes of maternal depression, from the child’s birth up to age nine, taking on values from 0 to 7. In Equation (2), the outcome *HK*_*Ct*_ is successively different measures of child *C*’s human capital at age *t*: the fine-grading average Key Stage test-scores at ages 11 and 14, the total GCSE score at age 16, and total (carer-reported) SDQ at child ages 11, 13 and 16. We standardise the different *HK*_*Ct*_ variables for comparison purposes, as they are not measured on the same scale.

We address pleiotropy by controlling for a set of both mother and child traits. *X*_*M*_ is a vector of mother’s traits when the child is aged nine: age at the birth of the child and dummies for being employed, having at least an A-level,^8^ having a partner, having a partner with at least an A-level, having an employed partner, the number of additional children, and banded household income (the latter two variables are measured at child age 8). It can be argued that some of these are potentially bad controls, as they may themselves be influenced by maternal depression (for example, mother’s labour-force status and the household’s income). We will address this issue in the robustness checks. Last, the *X*_*C*_ are time-invariant child traits: gender, birth year and birth order.

In Equation (1), 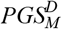, the maternal polygenic score for depression (our measure of mother’s genetic variants, 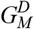, in Figure 1) is used as an instrument for maternal depression *D*_*M*_. We calculate 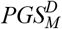 using the command-line program PLINK 1.9, with summary statistics from the single-trait depression meta-analysis GWAS in Turley *et al*. (2018). 68 of the 88 SNPs identified in the GWAS are genotyped in ALSPAC participants and were used in the PGS: see Appendix B for the details of the calculation. We standardise 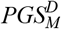, as polygenic scores have no natural scale. With polygenic scores being based on genetic variants that are determined at conception, the exogenous variation in maternal depression provided by 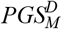 is fixed prior to the child’s birth: this rules out reverse-causality concerns (e.g. mothers’ mental health being affected by their children’s poor cognitive and/or non-cognitive performance).

We address population stratification by excluding mothers of non-European descent: Hansell *et al*. (2015) find no evidence of any remaining population stratification in ALSPAC after this selection and other standard quality-control (QC) procedures (see Taylor *et al*., 2018, for a complete overview of the QC procedures that were applied to ALSPAC data prior to its release). While the documented lack of stratification provides evidence in favour of the independence assumption in our context, we always control for 10 ancestry-informative principal components *PC*_*M*_ (as in von Hinke *et al*., 2016) and carry out additional tests for the influence of grandparental characteristics and partners’ depression on the effect of maternal depression (see Section 4.3).

Our estimation sample consists of observations with non-missing values for mothers’ genetic information, depression history, and the controls measured at child age nine.^9^ As there are only 1,065 families with non-missing information on all six human-capital measures and we cannot reject concerns about weak instruments in this balanced sample, we here use a different estimation sample for each dependent variable to maximise statistical power. Our final samples consist of between 2,036 and 2,993 observations per equation estimated. Due to attrition, the size of the non-cognitive skills estimation samples falls naturally with child age (from 2,993 to 2,076 observations). For cognitive skills, the estimation samples consist of 2,828 observations at age 16 (GCSE), 2,601 observations at age 11 (KS2) and 2,036 at age 14 (KS3). The discrepancy between the sample sizes at age 16 and earlier child ages reflects that the average KS2 and KS3 grades are retrospectively matched when the child takes her GCSE exams at age 16. 10% of the 227-observation difference between the GCSE and KS2 samples is due to either missing values in the school and academic year identifiers or in the grades, while the remaining 90% is due to the NPD data-cleaning process. For the gap between the GCSE and KS3 samples, 258 observations are missing for these two reasons, while the remaining 534 are due to the KS3 grades of ALSPAC children taking their GCSE in academic year 2008-09 no longer being collected.^10^ The influence of maternal depression and 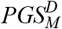 on attrition and the different sample sizes is discussed in the next section.

The distribution of self-reported maternal depression in the different estimation samples appears in Table 1, where depression takes on values between zero and seven. Around half of the women in our samples reported at least one episode of depression between the birth and ninth birthday of their child. This figure is consistent with data from a nationally-representative survey, the British Household Panel Study (BHPS), over the same time period: around 45% of the mothers observed for at least two consecutive years from 1991 to 2000 reported a least one episode of depression. The distribution of the measures of children’s human capital is shown in Appendix Figure A1, and the complete descriptive statistics are listed in Tables A2 (cognitive skills) and A3 (non-cognitive skills).

**Table 1:** The Distribution of Maternal Depression

|  | Estimation Sample: |  |  |  |  |  |
| --- | --- | --- | --- | --- | --- | --- |
|  | Test-scores |  |  | Total SDQ |  |  |
|  | Age 11<br>(1) | Age 14<br>(2) | Age 16<br>(3) | Age 11<br>(4) | Age 13<br>(5) | Age 16<br>(6) |
| Maternal Depression by Age 9: |  |  |  |  |  |  |
| No episodes | 47.1% | 47.4% | 47.0% | 48.6% | 50.3% | 50.5% |
| 1 episode | 17.0% | 16.6% | 17.2% | 16.7% | 16.8% | 17.1% |
| 2 episodes | 11.1% | 11.1% | 11.0% | 10.9% | 10.6% | 11.1% |
| 3 episodes | 8.0% | 7.5% | 7.7% | 7.2% | 7.1% | 6.7% |
| 4 episodes | 6.2% | 6.4% | 6.2% | 6.0% | 5.4% | 6.1% |
| 5 episodes | 4.5% | 4.6% | 4.5% | 4.4% | 4.2% | 3.7% |
| 6 episodes | 4.0% | 3.9% | 3.9% | 3.9% | 3.5% | 2.5% |
| 7 episodes | 2.4% | 2.6% | 2.4% | 2.4% | 2.1% | 2.3% |
| Observations | 2601 | 2036 | 2828 | 2993 | 2585 | 2076 |

*Note:* These figures refer to the estimation samples used in the main analysis.

While Equation (2) partly addresses pleiotropy by controlling for both the mother’s and the child’s traits (*X*_*M*_ and *X*_*C*_), the maternal PGS may still be directly linked to child human capital via the child’s genome (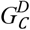 and 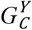 in Figure 1). We here follow Lawlor *et al*. (2017) and DiPrete *et al*. (2018), and address these concerns by controlling for the child’s PGS for depression and, respectively, cognitive and non-cognitive outcomes (see Section 4.2.3).

## 4. Results

### 4.1. Main Results

Table 2 presents the OLS and 2SLS estimates of Equation (2) for the effect of maternal depression on the different measures of child human capital. All of the estimated coefficients are negative and significantly different from zero at the 10% level at least. In the 2SLS results in columns (2), (4) and (6), one additional episode of maternal depression before child age nine reduces child test-scores by on average 23% of a standard-deviation (SD) and total SDQ by roughly 45% of a SD.^11^ Although the 2SLS estimates become a little larger as the child grows older, none of them are significantly different from each other. This pattern does not reflect the different sample compositions: restricting our analysis to families with valid information on either all of the cognitive-skill measures or all of the non-cognitive skill measures yields similar conclusions (these results are available upon request). Our specification exploits the longitudinal dimension of the dataset by looking at the impact of the observed history of a mother’s depression on the subsequent cognitive and non-cognitive outcomes of her children. Our estimates may thus reflect the predictive effect of the PGS on unobserved later episodes of maternal depression occurring between child age nine and the time the child’s outcome of interest is observed. Maternal depression during a child’s puberty could have a greater impact on their schoolwork and behaviour, producing larger coefficients at ages 14 and 16. In either case, maternal depression produces worse child outcomes.

**Table 2:** Maternal Depression and Child Human Capital: OLS and 2SLS Results

|  | Test-scores |  |  |  |  |  |
| --- | --- | --- | --- | --- | --- | --- |
|  | Age 11 |  | Age 14 |  | Age 16 |  |
|  | OLS<br>(1) | 2SLS<br>(2) | OLS<br>(3) | 2SLS<br>(4) | OLS<br>(5) | 2SLS<br>(6) |
| Maternal Depression | -0.018*<br>(0.010) | -0.222*<br>(0.120) | -0.031***<br>(0.010) | -0.178*<br>(0.097) | -0.016*<br>(0.009) | -0.273***<br>(0.102) |
| *First Stage:* |  |  |  |  |  |  |
| Mother's PGS |  | 0.158***<br>(0.042) |  | 0.205***<br>(0.046) |  | 0.179***<br>(0.039) |
| F-statistics |  | 14.1 |  | 19.5 |  | 21.2 |
| Observations | 2601 | 2601 | 2036 | 2036 | 2828 | 2828 |
|  | Total SDQ |  |  |  |  |  |
|  | Age 11 |  | Age 13 |  | Age 16 |  |
|  | OLS<br>(1) | 2SLS<br>(2) | OLS<br>(3) | 2SLS<br>(4) | OLS<br>(5) | 2SLS<br>(6) |
| Maternal Depression | -0.100***<br>(0.009) | -0.390***<br>(0.128) | -0.087***<br>(0.010) | -0.421***<br>(0.136) | -0.083***<br>(0.012) | -0.531**<br>(0.245) |
| *First Stage:* |  |  |  |  |  |  |
| Mother's PGS |  | 0.160***<br>(0.035) |  | 0.167***<br>(0.037) |  | 0.114***<br>(0.040) |
| F-statistics |  | 20.9 |  | 20.8 |  | 8.1 |
| Observations | 2993 | 2993 | 2585 | 2585 | 2076 | 2076 |

Notes: Standard errors appear in parentheses. All dependent variables are standardised. Maternal depression is the number of episodes of depression reported by the mother from the birth of the child to the child's ninth birthday. The controls are: the child's gender, birth year, birth order, number of siblings, the age of the mother at the birth of the child; dummies for the mother having at least an A-level, being employed, having a partner, having a partner with at least an A-level, having an employed partner, dummies for banded household income and ancestry-informative principal components. All regressions using test-scores as the dependent variable include school and school-year fixed effects. The reported F-statistics are those for the Cragg-Donald Wald weak-identification test. \*\*\* p<0.01, \*\* p<0.05, \* p<0.1.

Instrument relevance is evaluated in the first-stage estimates below the 2SLS results in Table 2. As expected, a higher PGS for depression significantly predicts more maternal-depression episodes in all specifications (at the 0.1% level at least). We also list the Cragg-Donald Wald F-statistics for the first-stages, which are sufficiently large to alleviate weak-instrument concerns in most cases.^12^ This F-statistic is under 10 only for the effect of maternal depression on total SDQ at age 16, which may show selective attrition. The probability of dropping out of the total SDQ estimation sample between two periods rises with maternal depression, but does not depend on the value of the instrument. It is thus unsurprising to see a lower first-stage F-statistic in the last column of the bottom panel of Table 2.^13^

Columns (1), (3) and (5) show the OLS results. Although these are qualitatively similar to the 2SLS estimates, they are four to ten times smaller in size. This gap may reflect that the GWAS summary statistics from Turley *et al*. (2018) are based on discovery samples where the trait is mostly measured as clinically diagnosed depression or self-diagnosed major depressive disorder (in around 80% of cases). As such, it is normal that the 2SLS estimates be larger than those in OLS, as the instrument captures more extreme forms of depression, that in turn play a larger role in human-capital accumulation. When analysing a non-binary trait (like our measure of maternal depression) genetic compliers can be seen as the whole population (see Dixon *et al*., 2020). Our 2SLS estimates then capture the average treatment effect of the trait that is most prevalent in the GWAS discovery cohorts – that is, ‘severe’ forms of depression (clinically-diagnosed depression, or major depressive disorder). On the contrary, the OLS estimates reveal the average effect of all forms of depression, both mild and severe.

The difference between the OLS and 2SLS estimates is larger for cognitive than non-cognitive skills: this may reflect the relative importance of diagnosed and undiagnosed symptoms of maternal depression in these two dimensions of human capital. While we do not observe formal diagnoses of depression, we know whether the mother consulted a doctor due to her depressive symptoms. When we separately consider episodes of maternal depression that were followed up by a medical visit and those that were not, the descriptive evidence from the OLS estimates suggests that, while both measures matter equally for non-cognitive skills, only the former is significantly associated with child cognitive skills (results available upon request).^14^

### 4.2. Addressing the Exclusion Restriction

#### 4.2.1. Horizontal Pleiotropy

The credibility of the exclusion restriction relies on there being no relationship between the PGS for maternal depression and the child outcomes, other than via maternal depression. However, as set out in Section 3.2, a genetic variant may predict more than one trait: this is horizontal pleiotropy. While we already control for a set of maternal traits in our main specification, we here provide additional evidence against pleiotropy playing a significant role in our analysis. Table A4 in Appendix A shows the bivariate associations between the PGS for depression and a variety of maternal traits. Unsurprisingly, the association between the PGS and maternal depression is positive and very significant. Just as importantly, none of the other traits is significantly associated with this instrument. While we cannot entirely rule out an effect of the genetic variants in the mother’s PGS on other unobserved traits involved in child human-capital development, the lack of any correlation with the observed traits is reassuring.

We also address the risk of pleiotropy more directly, investigating the known biological functions that are linked to the 68 SNPs used in the mother’s PGS for depression. We do so using the NHGRI-EBI online GWAS Catalog to review all of the biological functions associated with our SNPs. In line with von Hinke *et al*. (2016), we then calculate a new PGS discarding the six lead SNPs linked to either the cognitive or non-cognitive outcomes,^15^ as these are likely to violate the exclusion restriction via their effect on the mother’s human capital. Columns (2), (4) and (6) of Table A5 list the 2SLS estimates with this restricted PGS: these are very similar to those in the baseline (reproduced in columns (1), (3) and (5)). We also calculate the mother’s PGS for depression excluding the lead SNPs that predict any trait other than depression, even those that may appear unrelated to human capital (e.g. bone density). The last two sets of mother’s PGS exclude the SNPs in LD with genetic variants explaining other traits (first, only the cognitive and/or non-cognitive outcomes, and second an expanded set of traits made up of these two outcomes, along with BMI, and smoking), using a window of 500k base-pairs and a squared pairwise correlation of at least 0.6. Although both approaches reduce the variability in our instrument on which identification is based, the 2SLS estimates remain qualitatively the same. These results are available upon request.

We last address unobserved associations between the SNPs for depression and mother’s human capital (e.g. unknown biological pathways) by computing her PGS for both cognitive and non-cognitive skills based on the GWAS summary statistics in Demange *et al*. (2021), and introducing these as controls in our main specification. Table A6 shows that partialling out maternal genetic variation in cognitive and non-cognitive skills does not qualitatively change the results (and these latter are mostly not significant predictors of maternal depression in the first-stage regressions).

#### 4.2.2. Bad Controls

As discussed in Section 2, controlling for mother’s and child’s traits attenuates pleiotropy concerns. It can nonetheless be argued that some of these traits (for example, mother’s labour-force status, the presence of a partner in the household and household income) are bad controls as they could themselves result from depression. We thus re-estimate our 2SLS regressions first with no controls, then controlling for the mother’s traits, and finally for the child’s traits. The results, as compared to the baseline estimates (which control for both sets of traits), are depicted in Figure 2. The inclusion of potentially ‘bad’ controls makes relatively little difference, and the estimated coefficients on maternal depression remain negative and significant in every specification for every outcome.

**Figure 2:**
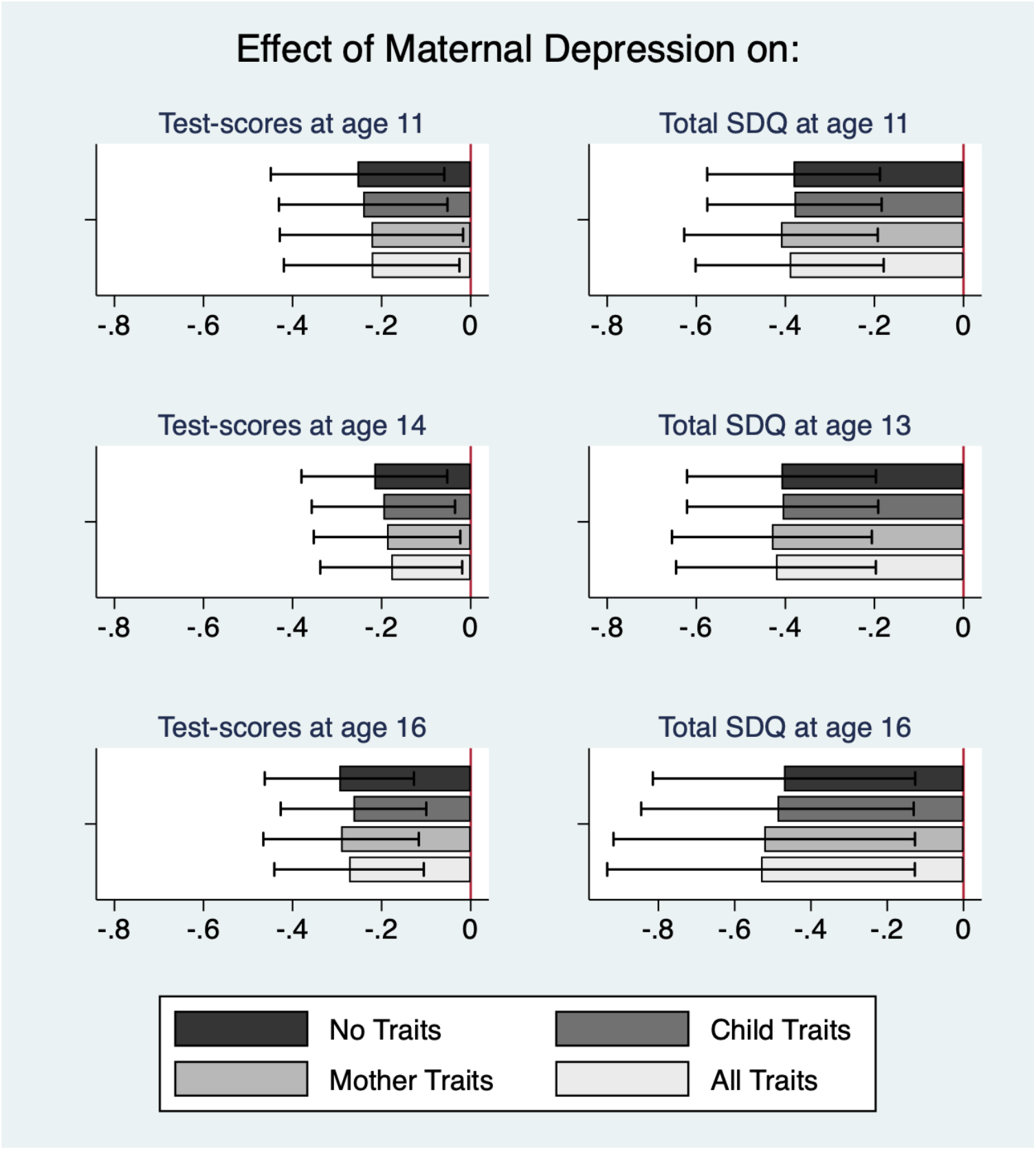
Maternal Depression and Child Human Capital: 2SLS Results with Different Sets of Covariates *Notes*: The horizontal lines in each bar show the 90% confidence intervals. All of the dependent variables are standardised. The child traits are the child’s gender, birth year and birth-order dummies. The mother’s traits are the child’s number of siblings in the household, the age of the mother at birth of the cohort member, dummies for the mother having at least an A-level, working, having a partner, having a partner with at least an A-level, and having a working partner, and dummies for banded household income. All regressions using test-scores as the dependent variable include school and school-year fixed effects. The Cragg-Donald Wald F-statistics for weak identification, going from the “No traits” specification to the “All traits” specification, are the following: for KS2, 15.5, 16.0, 13.0, 14.0; for KS3, 20.9, 20.7, 18.8, 19.5; for KS4, 22.8, 22.2, 20.6, 21.2; for SDQ11, 23.8, 23.0, 20.6, 21.0; for SDQ13, 22.1, 21.4, 21.1, 20.8; for SDQ16, 9.7, 9.2, 8.3, 8.1. ^***^ p<0.01, ^**^ p<0.05, ^*^ p<0.1.

#### 4.2.3. Genetic Inheritance and Trait Overlap

We can expect about half of the genetic variants included in the PGS for maternal depression to be passed on to the child (and an even higher figure if the parents match assortatively on the basis of depression). As noted in Section 2, if the inherited variants are correlated with the child’s cognitive/non-cognitive outcomes, then the exclusion restriction will be violated. Controlling for the child’s polygenic scores for depression and cognitive/non-cognitive skills will effectively shut off any confounding effect from genetic inheritance that affects these traits.

We here again use the summary statistics from the depression meta-analysis GWAS in Turley *et al*. (2018) to calculate a PGS for depression in children. We use the GWAS-by-subtraction summary data from Demange *et al*. (2021) and the summary statistics from the genome-wide association meta-analysis of Middeldorp *et al*. (2016) to calculate the PGSs for cognitive and non-cognitive skills. We do not use Demange *et al*. (2021) to calculate the PGS for non-cognitive skills, as the weights from a GWAS on an adult population might not be relevant for children (see Zhang *et al*., 2015). Furthermore, as Demange *et al*. (2021) define as ‘non-cognitive’ those SNPs associated with educational attainment independent of cognitive ability, a PGS based on their summary statistics may not be appropriate for our measure of non-cognitive skills (SDQ). In contrast, Middeldorp *et al*. (2016) use a discovery sample of children under 13 to identify the SNPs associated with attention-deficit/hyperactivity disorder (ADHD) symptoms (a condition arguably captured by the ‘inattention/hyperactivity’ subscale of the SDQ).^16^

Columns (1), (5) and (9) in Table 3 show the baseline 2SLS estimated coefficients for maternal depression from Table 2; the other columns introduce various child PGS measures. As the child genotype is missing in roughly 10% of the cases, we replace the missing values with the sample average and use a missing-indicator flag (dropping missing-genotype children from the estimation produces similar results). Columns (2), (6) and (10) in the top panel of Table 3 control for the child’s cognitive-skill PGS: this is positively correlated with the child’s average test-scores (as expected), but not with the PGS for maternal depression (there is little change in the F-statistics). Analogous results pertain for the child’s non-cognitive PGS in the bottom panel of Table 3.

**Table 3:** Addressing Genetic Inheritance for Maternal Depression and Child Human Capital: 2SLS results

|  | Test-scores |  |  |  |  |  |  |  |  |  |  |  |
| --- | --- | --- | --- | --- | --- | --- | --- | --- | --- | --- | --- | --- |
|  | Age 11 |  |  |  | Age 14 |  |  |  | Age 16 |  |  |  |
|  | (1) | (2) | (3) | (4) | (5) | (6) | (7) | (8) | (9) | (10) | (11) | (12) |
| Maternal Depression | -0.222* | -0.192 | -0.238* | -0.225* | -0.178* | -0.164* | -0.182** | -0.177** | -0.273*** | -0.255** | -0.294*** | -0.288*** |
|  | (0.120) | (0.117) | (0.123) | (0.121) | (0.097) | (0.096) | (0.089) | (0.089) | (0.102) | (0.100) | (0.106) | (0.105) |
| Child PGS:<br>Cognitive Skills |  | 0.071*** |  | 0.072*** |  | 0.047** |  | 0.047** |  | 0.061*** |  | 0.062*** |
|  |  | (0.020) |  | (0.021) |  | (0.022) |  | (0.022) |  | (0.020) |  | (0.020) |
| Child PGS:<br>Depression |  |  | 0.005 | 0.012 |  |  | 0.002 | 0.006 |  |  | 0.009 | 0.014 |
|  |  |  | (0.022) | (0.021) |  |  | (0.021) | (0.021) |  |  | (0.021) | (0.021) |
| F-statistics | 14.1 | 13.9 | 13.7 | 13.7 | 19.5 | 19.2 | 23.0 | 22.9 | 21.2 | 21.3 | 20.3 | 20.4 |
| Observations | 2601 | 2601 | 2601 | 2601 | 2036 | 2036 | 2036 | 2036 | 2828 | 2828 | 2828 | 2828 |
|  | Total SDQ |  |  |  |  |  |  |  |  |  |  |  |
|  | Age 11 |  |  |  | Age 13 |  |  |  | Age 16 |  |  |  |
|  | (1) | (2) | (3) | (4) | (5) | (6) | (7) | (8) | (9) | (10) | (11) | (12) |
| Maternal Depression | -0.390*** | -0.391*** | -0.295** | -0.296** | -0.421*** | -0.421*** | -0.384*** | -0.383*** | -0.531** | -0.529** | -0.478* | -0.482* |
|  | (0.128) | (0.128) | (0.124) | (0.124) | (0.136) | (0.136) | (0.127) | (0.127) | (0.245) | (0.244) | (0.249) | (0.250) |
| Child PGS:<br>Non-Cognitive Skills |  | 0.044** |  | 0.039* |  | 0.058** |  | 0.056** |  | 0.077** |  | 0.074** |
|  |  | (0.022) |  | (0.021) |  | (0.025) |  | (0.024) |  | (0.031) |  | (0.031) |
| Child PGS:<br>Depression |  |  | -0.036* | -0.036* |  |  | -0.015 | -0.015 |  |  | -0.014 | -0.013 |
|  |  |  | (0.022) | (0.022) |  |  | (0.024) | (0.024) |  |  | (0.031) | (0.031) |
| F-statistics | 20.9 | 20.9 | 19.5 | 19.4 | 20.8 | 20.8 | 22.4 | 22.4 | 8.1 | 8.1 | 7.1 | 7.0 |
| Observations | 2993 | 2993 | 2993 | 2993 | 2585 | 2585 | 2585 | 2585 | 2076 | 2076 | 2076 | 2076 |

Notes: Standard errors are in parentheses. All dependent variables are standardised. Maternal depression is the number of episodes of depression reported by the mother from the birth of the child to the child's ninth birthday. The controls are: the child's gender, birth year, birth order, number of siblings, the age of the mother at the birth of the child; dummies for the mother having at least an A-level, being employed, having a partner, having a partner with at least an A-level, having an employed partner, dummies for banded household income and ancestry-informative principal components. All regressions using test-scores as the dependent variable include school and school-year fixed effects. The reported F-statistics are those for the Cragg-Donald Wald weak-identification test. \*\*\* p<0.01, \*\* p<0.05, \* p<0.1.

We then turn to the child’s depression PGS, part of which is inherited from the mother. As expected, we find a 50% unconditional correlation between the mother’s and the child’s PGSs for depression (which explains the lower F-statistics in columns (3), (7) and (11) when controlling for the latter). However, in regressions including the mother’s PGS, the child’s depression PGS does not significantly influence the dependent variables (as shown in the table) or maternal depression (not reported – results available upon request). As the PGS uses weights derived from an adult population, the genetic variants identified there may not work in the same way for children.

Columns (4), (8) and (12) introduce the two scores simultaneously, which does not change our conclusions: the children of more-depressed mothers have significantly worse cognitive and non-cognitive skills. Although the estimated maternal-depression coefficients change a little in size as we introduce different PGS controls, they are never significantly different from each other.^17^

#### 4.2.4. Plausible Exogeneity

While the analyses above have put considerable effort into tackling potential violations of the exclusion restriction, there may still be unobserved pathways for which we do not control. For instance, while we do account for horizontal pleiotropy from the mother’s genetic variants by controlling for a set of maternal covariates, there are still channels we do not observe or, if observed, are subject to measurement error and reporting bias. Additionally, although their impact is likely to be marginal, there might be yet some other sources of pleiotropy confounding our main estimates (see, for instance, network pleiotropy in Boyle *et al*., 2017).

We thus follow the analysis in Conley *et al*. (2012), and consider the implications of our instrument being only ‘plausibly exogenous’. Here the instrumental variable is allowed to have a direct effect, *λ*, on the outcomes. As in Nybom (2017), *λ* is the share of the reduced-form effect of the instrument on child human capital that is independent of the variable we instrument, maternal depression. Considering different values of *λ* allows us to identify the threshold at which our 2SLS estimated coefficients are no longer significant at the 10% level.

Figure A2 depicts the 2SLS estimates from Equation (2) for *λ* in the interval [0, 1]. We follow Nybom (2017) and assume that *λ* is known with certainty. For cognitive skills at ages 11 and 14, once *λ* reaches 0.1 the 2SLS estimates are no longer significant at the 10% level (as revealed by the grey shaded areas). For all other outcomes, the threshold is larger (*λ* from 0.3 up to 0.5). In other words, as long as the direct effect of the PGS for maternal depression on the child outcomes is under 30% of the total reduced-form effect, most of our 2SLS estimates remain significantly different from zero at the 10% level.^18^

### 4.3. The Influence of Maternal Grand-parents and the Partner

Based on the ethnic composition of our subsample of ALSPAC participants and the fact that we always control for 10 ancestry-informative principal components, we have little reason to believe that residual population stratification is a threat to the independence assumption (see Section 3.3). However, other concerns regarding the independence assumption remain. Mendel’s laws of Segregation and Independent Assortment imply that, conditional on the parental genotype, the child’s genotype is the result of a lottery. The genotypes of the maternal grandparents are not available in ALSPAC, so that the mother’s genotype, and consequently her PGS for depression, might partly capture the effect of her parents’ genotypes, with the latter also potentially being correlated with the *U* variables in Figure 1 (see Section 3.2).

While we cannot control for the genetic variants of the maternal grandparents, we do have data on a set of grandparental traits: their education, social status, and a dummy for at least one of the maternal grandparents having had a severe mental illness prior to the birth of the child. The results controlling for these variables appear in Table A7. The 2SLS estimates are virtually unchanged from those in the baseline. The F-statistics are slightly lower. This is unsurprising: even though, after conditioning on the mother’s traits, none of the grandparental characteristics is correlated with child human-capital, the mother having at least one parent with a history of mental illness is positively and significantly associated with both our measure of maternal depression and her PGS for depression.

We finally consider assortative matching between the child’s parents: depressed mothers might choose their partners according to certain traits (depression itself, and/or other traits), which may in turn affect child human capital. Our main specification, which includes a number of the mother’s partner’s controls, partly addresses this. We can further show that these traits (having a partner, partner’s working status and education) are not systematically explained by the mother’s PGS for depression (see Table A4). While this alleviates concerns about cross-trait assortative matching, mothers with a higher genetic risk of being depressed might be more likely to have a depressed partner. We have information on the mother’s partner’s number of depression episodes, measured at child ages 2, 4 and 6. While the unconditional correlation between the partner’s depression and maternal depression is relatively high (0.44) and significant, its correlation with the PGS for maternal depression is not statistically different from zero (in both bivariate and multivariate analyses). Introducing partner’s depression makes little difference to our main results: see Table A8.

### 4.4. Robustness Checks

#### 4.4.1. The Measurement of Maternal Depression

We carry out a battery of robustness checks. We first show that our results hold with different maternal-depression measures (the descriptive statistics of which appear in Table A9). Our baseline count of reported depressive episodes between child ages 0 and 9 weights recent and more-distant episodes equally, but those at younger child ages may matter more (as children then have greater developmental plasticity and spend more time with their mothers). Panels B and C of Table A10 however reveal larger estimates for more-recent depressive episodes (although the estimated coefficients between these panels are not significantly different from each other). The results continue to hold using only the number of episodes net of post-partum depression (i.e. between child ages 2 and 9) in Panel D, and with a dummy for any episode of depression in Panel E. Panel F considers a dummy for recent depression and Panel G the average of the six maternal scores on the Edinburgh Postnatal Depression Scale (EPDS) between child ages 0 and 8 (at child age 8 months and 2, 3, 5, 6 and 8 years). Although the results continue to be of the same nature, the F-statistics are notably worse. The instrument weakness here reveals that our PGS has greater predictive power when maternal depression is measured over longer time periods and in a similar way to that in the GWAS meta-analysis (the EPDS does not appear in Turley *et al*., 2018).

#### 4.4.2. The Measurement of Non-Cognitive Skills

The SDQ measure of non-cognitive skills we use is reported by the mother. As depressed mothers may over- or under-estimate their children’s non-cognitive skills (Del Bono *et al*., 2020) we turn to teacher-reported SDQ (which is only available when the child was aged 11). In the first column of Table A11, an additional episode of maternal depression continues to reduce total SDQ with an effect size identical to that in Table 2.^19^ We also test for convergent validity using the SMFQ (reported by the main carer) in columns (2) to (4) of Table A11: the resulting estimates are not significantly different from those in the baseline (although that at age 16 is statistically insignificant).

## 5. Conclusion

Social scientists are interested in causal phenomena, and research agendas are partly limited to the analysis of variables that can be influenced, either directly or via policy intervention. However, there are many variables and pathways that are either costly or impossible to manipulate. We believe that it is possible to make causal statements about some of these latter via the increasing availability of genetic data and recent developments in the fields of Epidemiology and Molecular Genetics. This is the approach that we have taken here. However, the use of genetic data as instruments is not a quick fix, as it comes with a number of quite-stringent assumptions. We have here discussed a number of tests and tools that can be applied in this empirical setting.

We illustrate how genetic data can be used to identify the effect of maternal depression on children’s human capital, using data from a British birth-cohort study. We first show that genetic variants, combined into a synthetic polygenic score, are a strong instrumental variable for maternal depression. In 2SLS estimation, we then exploit the exogenous differences in maternal depression resulting from the mother’s genes to identify its negative consequences on the cognitive and non-cognitive outcomes of their adolescent children.

Our results suggest that fewer episodes of maternal depression will not only benefit mothers, but also improve their children’s human capital. In turn, better cognitive and non-cognitive skills in childhood are known to have positive returns on a variety of outcomes during adulthood, such as income and labour-market experience (Heckman *et al*., 2006; Heckman *et al*., 2018; Clark *et al*., 2018; Clark and Lepinteur, 2019). As revealed by the evaluation of the Improving Access to Psychological Therapies programme in the UK in Clark (2018), the costs of effective treatments for depression are extremely low compared to their expected benefits. If treatment also produces positive spillovers on children, the benefit-cost ratio will be even higher, making treatment more attractive.

However, as we compare depressed to not-depressed or less-depressed mothers using cross-section data on adolescents, our results do not tell us how changes in depression (in particular, due to its treatment) would affect children. Baranov *et al*. (2020) find only small long-term effects on child development following the treatment of prenatally-depressed mothers in rural Pakistan. The socio-economic, geographical and temporal contexts of our work and those in Baranov *et al*. (2020) are of course dissimilar. More importantly, they look at mothers who were already depressed pre-birth, whereas we consider a general sample of mothers, some of whom experience episodes of depression after birth and some of whom do not. While we show that the experience of maternal depression has large scarring effects on adolescent children, we do not know how easy it is to erase these scars. Policies that aim to prevent depression, rather than treat it once it occurs, may have a greater return from a societal perspective.

The use of polygenic scores as instrumental variables is a promising avenue for causal inference in observational data. It is however important to keep in mind that the genetic component of complex traits, such as mental health, is far from deterministic. The same polygenic score can be found in individuals with a very wide range of values of the trait of interest. This may reflect that the individual genetic architecture predicts outcomes partly via individuals’ reactions to their environment. This opens the door to policy intervention: while genes are fixed, the environment is not. Future research on which stressors are the most important in this context will help advance our understanding of the sign and size of causal relationships that can serve as inputs to public-policy debate.

## Data Availability

Data is available through application to the research executive of ALSPAC. Please note that the study website contains details of all the data that is available through a fully searchable data dictionary and variable search tool: http://www.bristol.ac.uk/alspac/researchers/our-data/.

## Appendix A: Additional Figures and Tables

**Figure A1:**
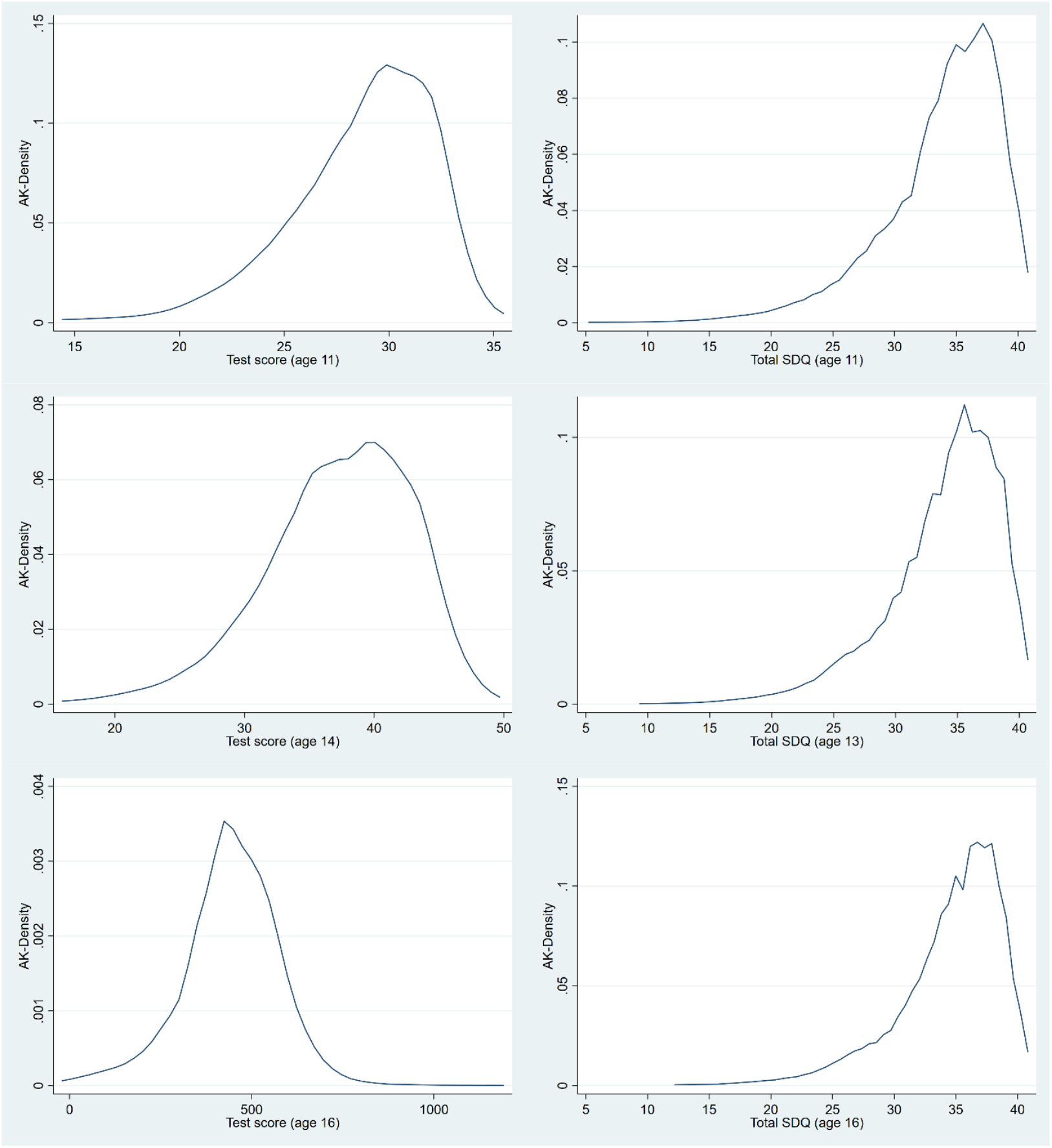
The Distribution of Test-scores and Total SDQ *Notes*: Each figure refers to one of our estimation samples. The test-scores at age 11, 14 and 16 respectively refer to the Key-Stage 2 average score, Key-Stage 3 average score and GCSE total score. The densities are plotted using an adaptive-kernel (see Van Kerm, 2003, for the technical details).

**Figure A2:**
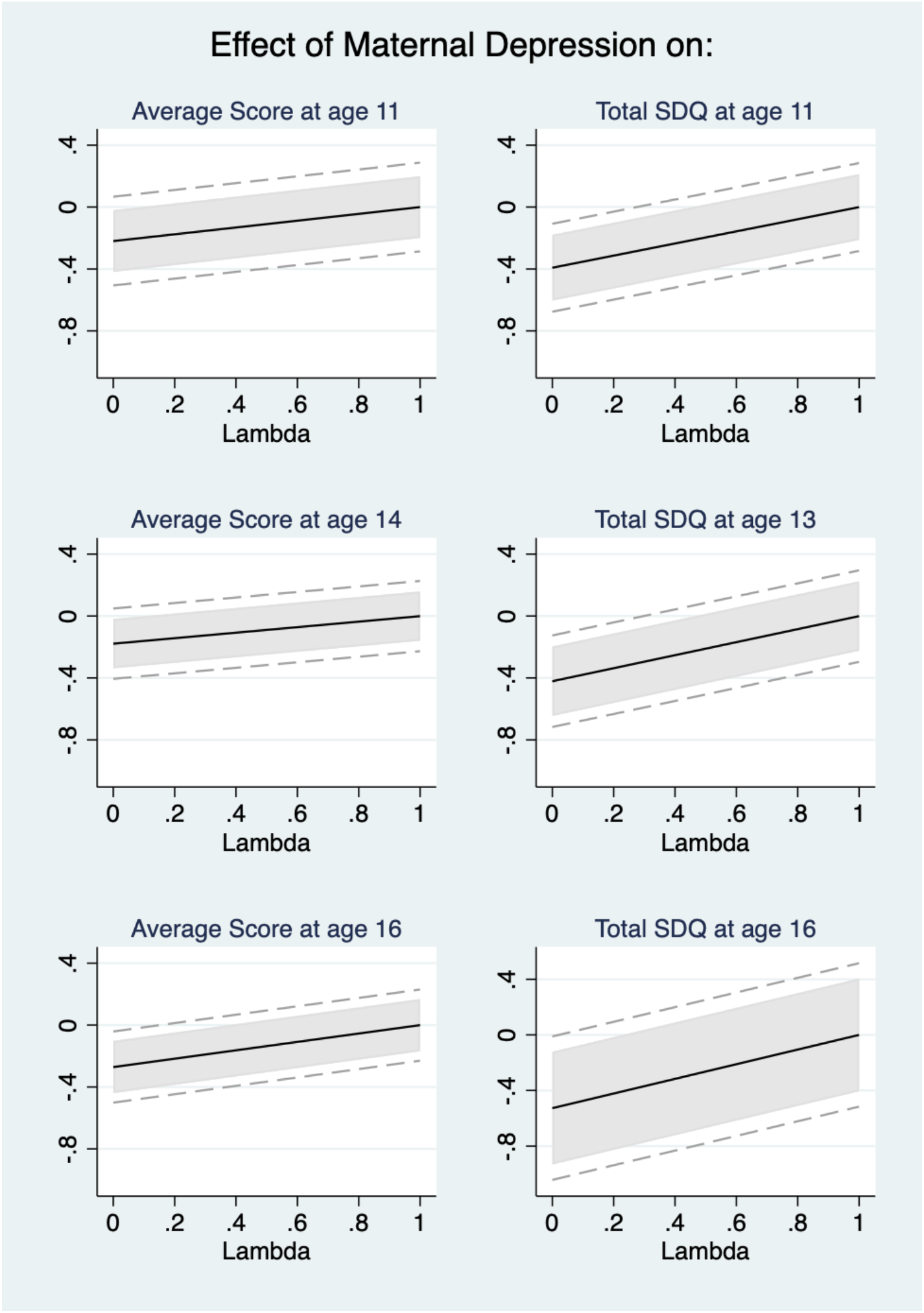
Plausible Exogeneity and Pleiotropy-robust MR *Notes*: Lambda represents the share of the reduced-form effect of the instrument on the outcome that is independent of maternal depression. The black line is the 2SLS point estimate of maternal depression for different values of lambda; the grey solid area represents 90% confidence intervals using the Nybom (2017) approach, while the grey dashed lines are the 90% confidence intervals following van Kippersluis and Rietveld (2018).

**Table A1:** Strengths and Difficulties Questionnaire (SDQ) *Please think about this child’s behaviour over the last 6 months if you can*

| This child: | Not True | Somewhat True | Certainly True |
| --- | --- | --- | --- |
| 1. Is considerate of other people's feelings [PS] | 2 | 1 | 0 |
| 2. Is restless, overactive, cannot stay still for long [H] | 0 | 1 | 2 |
| 3. Often complains of headaches, stomach-aches or sickness [E] | 0 | 1 | 2 |
| 4. Shares readily with other children, for example toys, treats, pencils [PS] | 2 | 1 | 0 |
| 5. Often loses temper [B] | 0 | 1 | 2 |
| 6. Is rather solitary, prefers to play alone [P] | 0 | 1 | 2 |
| 7. Is generally well behaved, usually does what adults request [B] | 2 | 1 | 0 |
| 8. Has many worries or often seems worried [E] | 0 | 1 | 2 |
| 9. Is helpful if someone is hurt, upset or feeling ill [PS] | 2 | 1 | 0 |
| 10. Is constantly fidgeting or squirming [H] | 0 | 1 | 2 |
| 11. Has at least one good friend [P] | 2 | 1 | 0 |
| 12. Often fights with other children or bullies them [B] | 0 | 1 | 2 |
| 13. Is often unhappy, depressed or tearful [E] | 0 | 1 | 2 |
| 14. Is generally liked by other children [P] | 2 | 1 | 0 |
| 15. Is easily distracted, concentration wanders [H] | 0 | 1 | 2 |
| 16. Is nervous or clingy in new situations, easily loses confidence [E] | 0 | 1 | 2 |
| 17. Is kind to younger children [PS] | 2 | 1 | 0 |
| 18. Often lies or cheats [B] | 0 | 1 | 2 |
| 19. Is picked on or bullied by other children [P] | 0 | 1 | 2 |
| 20. Often volunteers to help others (parents, teachers, other children) [PS] | 2 | 1 | 0 |
| 21. Thinks things out before acting [H] | 2 | 1 | 0 |
| 22. Steals from home, school or elsewhere [B] | 0 | 1 | 2 |
| 23. Gets along better with adults than with other children [P] | 0 | 1 | 2 |
| 24. Has many fears, is easily scared [E] | 0 | 1 | 2 |
| 25. Has a good attention span, sees chores or homework through to the end [H] | 2 | 1 | 0 |

*Notes:* [E], [B], [H], [P] and [PS] refer respectively to the following dimensions of the SDQ: "Emotional health", "Behaviour problems", "Hyperactivity/inattention", "Peer relationship problems" and "Pro-social behaviour". Internalising SDQ is the sum of the "Emotional health" and "Peer relationship problems" scores, and externalising SDQ the sum of the "Hyperactivity/inattention" and "Behaviour problems" scores. Total SDQ is the sum of internalising and externalising SDQ.

**Table A2:** Descriptive Statistics: Cognitive-Skills Estimation Samples

|  | Estimation Samples: |  |  |  |  |  |  |  |  |  |  |  |
| --- | --- | --- | --- | --- | --- | --- | --- | --- | --- | --- | --- | --- |
|  | Test-scores (age 11)<br>2601 observations |  |  |  | Test-scores (age 14)<br>2036 observations |  |  |  | Test-scores (age 16)<br>2828 observations |  |  |  |
|  | Mean | SD | Min | Max | Mean | SD | Min | Max | Mean | SD | Min | Max |
| *Cohort member characteristics:* |  |  |  |  |  |  |  |  |  |  |  |  |
| KS2 average score (fine-grading) | 28.76 | 3.48 | 15 | 35 |  |  |  |  |  |  |  |  |
| KS3 average score (fine-grading) |  |  |  |  | 37.33 | 5.65 | 17 | 49 |  |  |  |  |
| GCSE total score |  |  |  |  |  |  |  |  | 445.48 | 132.63 | 0 | 1171 |
| Female | 0.51 |  | 0 | 1 | 0.52 |  | 0 | 1 | 0.51 |  | 0 | 1 |
| Birth year: |  |  |  |  |  |  |  |  |  |  |  |  |
| 1991 | 0.36 |  | 0 | 1 | 0.43 |  | 0 | 1 | 0.36 |  | 0 | 1 |
| 1992 | 0.63 |  | 0 | 1 | 0.57 |  | 0 | 1 | 0.64 |  | 0 | 1 |
| 1993 | 0.01 |  | 0 | 1 | 0.00 |  | 0 | 0 | 0.01 |  | 0 | 1 |
| Birth order: |  |  |  |  |  |  |  |  |  |  |  |  |
| 1<sup>st</sup>-born | 0.46 |  | 0 | 1 | 0.45 |  | 0 | 1 | 0.46 |  | 0 | 1 |
| 2<sup>nd</sup>-born | 0.37 |  | 0 | 1 | 0.38 |  | 0 | 1 | 0.37 |  | 0 | 1 |
| 3<sup>rd</sup>-born | 0.14 |  | 0 | 1 | 0.13 |  | 0 | 1 | 0.14 |  | 0 | 1 |
| 4<sup>th</sup>-born or higher | 0.03 |  | 0 | 1 | 0.03 |  | 0 | 1 | 0.03 |  | 0 | 1 |
| *Mother and family characteristics:* |  |  |  |  |  |  |  |  |  |  |  |  |
| No. of episodes of maternal depression | 1.50 | 1.94 | 0 | 7 | 1.51 | 1.96 | 0 | 7 | 1.50 | 1.94 | 0 | 7 |
| Employed mother | 0.74 |  | 0 | 1 | 0.74 |  | 0 | 1 | 0.73 |  | 0 | 1 |
| Mother has an A-level or above | 0.33 |  | 0 | 1 | 0.33 |  | 0 | 1 | 0.35 |  | 0 | 1 |
| Age of the mother at child's birth | 29.29 | 4.23 | 18 | 44 | 29.29 | 4.23 | 18 | 44 | 29.43 | 4.28 | 18 | 44 |
| Presence of partner | 0.94 |  | 0 | 1 | 0.94 |  | 0 | 1 | 0.94 |  | 0 | 1 |
| Employed partner | 0.89 |  | 0 | 1 | 0.89 |  | 0 | 1 | 0.88 |  | 0 | 1 |
| Partner has an A-level or above | 0.47 |  | 0 | 1 | 0.47 |  | 0 | 1 | 0.49 |  | 0 | 1 |
| Number of siblings | 2.29 | 0.77 | 0 | 6 | 2.29 | 0.77 | 0 | 6 | 2.28 | 0.76 | 0 | 6 |
| Average family take-home income per week: |  |  |  |  |  |  |  |  |  |  |  |  |
| <£100 | 0.01 |  | 0 | 1 | 0.01 |  | 0 | 1 | 0.01 |  | 0 | 1 |
| £100-£199 | 0.07 |  | 0 | 1 | 0.07 |  | 0 | 1 | 0.07 |  | 0 | 1 |
| £200-£299 | 0.16 |  | 0 | 1 | 0.16 |  | 0 | 1 | 0.15 |  | 0 | 1 |
| £300-£399 | 0.20 |  | 0 | 1 | 0.21 |  | 0 | 1 | 0.20 |  | 0 | 1 |
| £400+ | 0.47 |  | 0 | 1 | 0.46 |  | 0 | 1 | 0.48 |  | 0 | 1 |
| Do not know | 0.05 |  | 0 | 1 | 0.05 |  | 0 | 1 | 0.05 |  | 0 | 1 |
| Missing | 0.04 |  | 0 | 1 | 0.04 |  | 0 | 1 | 0.04 |  | 0 | 1 |

Note: These figures refer to our estimation samples.

**Table A3:** Descriptive Statistics: Non-Cognitive Skills Estimation Samples

|  | Estimation Samples: |  |  |  |  |  |  |  |  |  |  |  |
| --- | --- | --- | --- | --- | --- | --- | --- | --- | --- | --- | --- | --- |
|  | Total SDQ (age 11) |  |  |  | Total SDQ (age 13) |  |  |  | Total SDQ (age 16) |  |  |  |
|  | 2993 observations |  |  |  | 2585 observations |  |  |  | 2076 observations |  |  |  |
|  | Mean | SD | Min | Max | Mean | SD | Min | Max | Mean | SD | Min | Max |
| *Cohort member characteristics:* |  |  |  |  |  |  |  |  |  |  |  |  |
| Total SDQ | 33.79 | 4.85 | 6 | 40 | 33.86 | 4.55 | 10 | 40 | 34.55 | 4.26 | 13 | 40 |
| Female | 0.50 |  | 0 | 1 | 0.50 |  | 0 | 1 | 0.53 |  | 0 | 1 |
| Birth year: |  |  |  |  |  |  |  |  |  |  |  |  |
| 1991 | 0.36 |  | 0 | 1 | 0.36 |  | 0 | 1 | 0.35 |  | 0 | 1 |
| 1992 | 0.63 |  | 0 | 1 | 0.63 |  | 0 | 1 | 0.64 |  | 0 | 1 |
| 1993 | 0.01 |  | 0 | 1 | 0.01 |  | 0 | 1 | 0.01 |  | 0 | 1 |
| Birth order: |  |  |  |  |  |  |  |  |  |  |  |  |
| 1<sup>st</sup>-born | 0.47 |  | 0 | 1 | 0.47 |  | 0 | 1 | 0.48 |  | 0 | 1 |
| 2<sup>nd</sup>-born | 0.37 |  | 0 | 1 | 0.37 |  | 0 | 1 | 0.37 |  | 0 | 1 |
| 3<sup>rd</sup>-born | 0.13 |  | 0 | 1 | 0.13 |  | 0 | 1 | 0.12 |  | 0 | 1 |
| 4<sup>th</sup>-born or more | 0.03 |  | 0 | 1 | 0.03 |  | 0 | 1 | 0.02 |  | 0 | 1 |
| *Mother and family characteristics:* |  |  |  |  |  |  |  |  |  |  |  |  |
| No. of episode of maternal depression | 1.46 | 1.94 | 0 | 7 | 1.37 | 1.88 | 0 | 7 | 1.33 | 1.84 | 0 | 7 |
| Employed mother | 0.73 |  | 0 | 1 | 0.74 |  | 0 | 1 | 0.74 |  | 0 | 1 |
| Mother has an A-level or more | 0.39 |  | 0 | 1 | 0.41 |  | 0 | 1 | 0.44 |  | 0 | 1 |
| Age of the mother at child's birth | 29.59 | 4.27 | 18 | 44 | 29.63 | 4.24 | 18 | 43 | 29.77 | 4.26 | 18 | 43 |
| Presence of partner | 0.94 |  | 0 | 1 | 0.94 |  | 0 | 1 | 0.95 |  | 0 | 1 |
| Employed partner | 0.89 |  | 0 | 1 | 0.89 |  | 0 | 1 | 0.90 |  | 0 | 1 |
| Partner has an A-level or above | 0.53 |  | 0 | 1 | 0.54 |  | 0 | 1 | 0.56 |  | 0 | 1 |
| Number of siblings | 2.27 | 0.76 | 0 | 6 | 2.26 | 0.75 | 0 | 6 | 2.25 | 0.74 | 0 | 6 |
| Average family take-home income per week: |  |  |  |  |  |  |  |  |  |  |  |  |
| <£100 | 0.01 |  | 0 | 1 | 0.01 |  | 0 | 1 | 0.01 |  | 0 | 1 |
| £100-£199 | 0.06 |  | 0 | 1 | 0.06 |  | 0 | 1 | 0.05 |  | 0 | 1 |
| £200-£299 | 0.14 |  | 0 | 1 | 0.13 |  | 0 | 1 | 0.13 |  | 0 | 1 |
| £300-£399 | 0.19 |  | 0 | 1 | 0.19 |  | 0 | 1 | 0.17 |  | 0 | 1 |
| £400+ | 0.51 |  | 0 | 1 | 0.53 |  | 0 | 1 | 0.55 |  | 0 | 1 |
| Do not know | 0.05 |  | 0 | 1 | 0.04 |  | 0 | 1 | 0.04 |  | 0 | 1 |
| Missing | 0.04 |  | 0 | 1 | 0.04 |  | 0 | 1 | 0.04 |  | 0 | 1 |

Note: These figures refer to our estimation samples.

**Table A4:** The PGS for Maternal Depression and Maternal Traits: Bivariate Associations

|  | PGS for Maternal Depression |
| --- | --- |
| *Maternal Traits:* |  |
| Maternal Depression [0-7] | 0.177***<br>(0.035) |
| Mother is Employed | -0.007<br>(0.008) |
| Mother has at least an A-level | 0.003<br>(0.009) |
| Age at Birth of CM | 0.066<br>(0.076) |
| Mother has a Partner | -0.005<br>(0.004) |
| Working Partner<sup>+</sup> | 0.002<br>(0.004) |
| Partner has at least an A-level | 0.012<br>(0.009) |
| Number of Siblings | 0.013<br>(0.014) |
| Family Income above the Median<sup>+</sup> | -0.013<br>(0.009) |

*Notes:* Each cell reports the estimate and standard errors (in parentheses) of separate bivariate associations between the PGS for maternal depression and various maternal traits. The bivariate regressions are based on the observations coming from our largest estimation sample (the one for which the SDQ at age 11 is not missing: 2993 observations). Using our five other estimation samples produces similar estimates.
<sup>+</sup> indicates that we excluded the missing values of the maternal traits from the estimation sample (including the missing values and introducing a missing-indicator flag produces similar estimates). \*\*\* p<0.01, \*\* p<0.05, \* p<0.1.

**Table A5:** Maternal Depression and Child Human Capital: 2SLS Results using the Mother’s PGS for Depression excluding Genetic Variants Linked to Known Biological Pathways

|  | Test-scores |  |  |  |  |  |
| --- | --- | --- | --- | --- | --- | --- |
|  | Age 11 |  | Age 14 |  | Age 16 |  |
|  | (1) | (2) | (3) | (4) | (5) | (6) |
| Maternal Depression | -0.222* | -0.214* | -0.178* | -0.142 | -0.273*** | -0.290*** |
|  | (0.120) | (0.124) | (0.097) | (0.103) | (0.102) | (0.110) |
| *First Stage:* |  |  |  |  |  |  |
| Mother's PGS for Depression | 0.158*** | 0.152*** | 0.205*** | 0.189*** | 0.179*** | 0.167*** |
|  | (0.042) | (0.042) | (0.046) | (0.047) | (0.039) | (0.039) |
| F-statistics | 14.1 | 13.0 | 19.5 | 16.5 | 21.2 | 18.7 |
| Observations | 2601 | 2601 | 2036 | 2036 | 2828 | 2828 |
|  | Total SDQ |  |  |  |  |  |
|  | Age 11 |  | Age 13 |  | Age 16 |  |
|  | (1) | (2) | (3) | (4) | (5) | (6) |
| Maternal Depression | -0.390*** | -0.396*** | -0.421*** | -0.418*** | -0.531** | -0.534* |
|  | (0.128) | (0.145) | (0.136) | (0.157) | (0.245) | (0.314) |
| *First Stage:* |  |  |  |  |  |  |
| Mother's PGS for Depression | 0.160*** | 0.142*** | 0.167*** | 0.145*** | 0.114*** | 0.090*** |
|  | (0.035) | (0.035) | (0.036) | (0.037) | (0.040) | (0.040) |
| F-statistics | 20.9 | 16.5 | 20.8 | 15.5 | 8.1 | 5.0 |
| Observations | 2993 | 2993 | 2585 | 2585 | 2076 | 2076 |

Notes: Standard errors appear in parentheses. All dependent variables are standardised. Maternal depression is the number of episodes of depression reported by the mother from the birth of the child to the child's ninth birthday. The controls are: the child's gender, birth year, birth order, number of siblings, the age of the mother at the birth of the child; dummies for the mother having at least an A-level, being employed, having a partner, having a partner with at least an A-level, having an employed partner, dummies for banded household income and ancestry-informative principal components. All regressions using test-scores as the dependent variable include school and school-year fixed effects. The reported F-statistics are those for the Cragg-Donald Wald weak-identification test. \*\*\* p<0.01, \*\* p<0.05, \* p<0.1.

**Table A6:** Maternal Depression and Child Human Capital: 2SLS Results Controlling for Mother’s PGS for Cognitive and Non-Cognitive Skills

|  | Test-scores |  |  |  |  |  |
| --- | --- | --- | --- | --- | --- | --- |
|  | Age 11 |  | Age 14 |  | Age 16 |  |
|  | (1) | (2) | (3) | (4) | (5) | (6) |
| Maternal Depression | -0.222* | -0.214* | -0.178* | -0.177* | -0.273*** | -0.270*** |
|  | (0.120) | (0.121) | (0.097) | (0.098) | (0.102) | (0.103) |
| *First Stage:* |  |  |  |  |  |  |
| Mother's PGS for Depression | 0.158*** | 0.156*** | 0.205*** | 0.202*** | 0.179*** | 0.177*** |
|  | (0.042) | (0.042) | (0.046) | (0.046) | (0.039) | (0.039) |
| Mother's PGS for Cognitive Skills |  | -0.049 |  | -0.093* |  | -0.030 |
|  |  | (0.046) |  | (0.050) |  | (0.042) |
| F-statistics | 14.1 | 13.5 | 19.5 | 18.9 | 21.2 | 20.8 |
| Observations | 2601 | 2601 | 2036 | 2036 | 2828 | 2828 |
|  | Total SDQ |  |  |  |  |  |
|  | Age 11 |  | Age 13 |  | Age 16 |  |
|  | (1) | (2) | (3) | (4) | (5) | (6) |
| Maternal Depression | -0.390*** | -0.392*** | -0.421*** | -0.422*** | -0.531** | -0.533** |
|  | (0.128) | (0.128) | (0.136) | (0.136) | (0.245) | (0.244) |
| *First Stage:* |  |  |  |  |  |  |
| Mother's PGS for Depression | 0.163*** | 0.164*** | 0.171*** | 0.172*** | 0.174*** | 0.129*** |
|  | (0.035) | (0.035) | (0.036) | (0.036) | (0.055) | (0.043) |
| Mother's PGS for Non-Cognitive Skills |  | -0.025 |  | -0.044 |  | -0.027 |
|  |  | (0.035) |  | (0.037) |  | (0.040) |
| F-statistics | 20.9 | 21.0 | 20.8 | 20.9 | 8.1 | 8.2 |
| Observations | 2993 | 2993 | 2585 | 2585 | 2076 | 2076 |

Notes: Standard errors appear in parentheses. All dependent variables are standardised. Maternal depression is the number of episodes of depression reported by the mother from the birth of the child to the child's ninth birthday. The controls are: the child's gender, birth year, birth order, number of siblings, the age of the mother at the birth of the child; dummies for the mother having at least an A-level, being employed, having a partner, having a partner with at least an A-level, having an employed partner, dummies for banded household income and ancestry-informative principal components. All regressions using test-scores as the dependent variable include school and school-year fixed effects. The reported F-statistics are those for the Cragg-Donald Wald weak-identification test. \*\*\* p<0.01, \*\* p<0.05, \* p<0.1.

**Table A7:** Maternal Depression and Child Human Capital: 2SLS Results Controlling for Grandparental Characteristics

|  | Test-scores |  |  | Total SDQ |  |  |
| --- | --- | --- | --- | --- | --- | --- |
|  | Age 11<br>(1) | Age 14<br>(2) | Age 16<br>(3) | Age 11<br>(4) | Age 13<br>(5) | Age 16<br>(6) |
| Maternal Depression | -0.255*<br>(0.132) | -0.186*<br>(0.100) | -0.296***<br>(0.112) | -0.385***<br>(0.138) | -0.437***<br>(0.147) | -0.525*<br>(0.273) |
| *First Stage:* |  |  |  |  |  |  |
| Mother's PGS | 0.146***<br>(0.042) | 0.196***<br>(0.047) | 0.164***<br>(0.039) | 0.147***<br>(0.035) | 0.158***<br>(0.037) | 0.102**<br>(0.040) |
| F-statistics | 11.8 | 17.5 | 17.7 | 17.6 | 18.2 | 6.4 |
| Observations | 2601 | 2036 | 2828 | 2993 | 2585 | 2076 |

Notes: Standard errors are in parentheses. All dependent variables are standardised. Maternal depression is the number of episodes of depression reported by the mother from the birth of the child to the child's ninth birthday. The controls are: the child's gender, birth year, birth order, number of siblings, the age of the mother at the birth of the child; dummies for the mother having at least an A-level, being employed, having a partner, having a partner with at least an A-level, having an employed partner, dummies for banded household income and ancestry-informative principal components. The regressions also include the following grandparental characteristics: dummies for the mother having at least one parent who had a mental illness before the birth of the child and the highest diploma obtained by the maternal grandmother and grandfather, and a social-status index for the maternal grandmother and grandfather. All of the regressions using test-scores as the dependent variable include school and school-year fixed effects. The reported F-statistics are those for the Cragg-Donald Wald weak-identification test. \*\*\* p<0.01, \*\* p<0.05, \* p<0.1.

**Table A8:** Maternal Depression and Child Human Capital: 2SLS Results Controlling for Partner’s Depression

|  | Test-scores |  |  | Total SDQ |  |  |
| --- | --- | --- | --- | --- | --- | --- |
|  | Age 11<br>(1) | Age 14<br>(2) | Age 16<br>(3) | Age 11<br>(4) | Age 13<br>(5) | Age 16<br>(6) |
| Maternal Depression | -0.240*<br>(0.129) | -0.187*<br>(0.102) | -0.297***<br>(0.113) | -0.406***<br>(0.136) | -0.436***<br>(0.143) | -0.566**<br>(0.270) |
| *First Stage:* |  |  |  |  |  |  |
| Mother's PGS | 0.148***<br>(0.040) | 0.195***<br>(0.044) | 0.163***<br>(0.037) | 0.151***<br>(0.033) | 0.160***<br>(0.035) | 0.105***<br>(0.039) |
| Partner's Depression | 0.965***<br>(0.068) | 0.963***<br>(0.073) | 0.942***<br>(0.062) | 0.912***<br>(0.056) | 0.859***<br>(0.060) | 0.873***<br>(0.066) |
| F-statistics | 13.4 | 19.3 | 19.3 | 20.3 | 20.4 | 7.4 |
| Observations | 2601 | 2036 | 2828 | 2993 | 2585 | 2076 |

Notes: Standard errors appear in parentheses. All dependent variables are standardised. Maternal depression is the number of episodes of depression reported by the mother from the birth of the child to the child's ninth birthday. The controls are: the child's gender, birth year, birth order, number of siblings, the age of the mother at the birth of the child; dummies for the mother having at least an A-level, being employed, having a partner, having a partner with at least an A-level, having an employed partner, dummies for banded household income and ancestry-informative principal components. All regressions using test-scores as the dependent variable include school and school-year fixed effects. The reported F-statistics are those for the Cragg-Donald Wald weak-identification test. \*\*\* p<0.01, \*\* p<0.05, \* p<0.1.

**Table A9:** Descriptive Statistics: Alternative Measures of Maternal Depression

|  | Mean | SD | Min | Max |
| --- | --- | --- | --- | --- |
| *Estimation Sample: Test-scores (age 11)* |  |  |  |  |
| No. of depressive episodes between child age 0 and 5 | 1.06 | 1.44 | 0 | 5 |
| No. of depressive episodes between child age 5 and 9 | 0.44 | 0.70 | 0 | 2 |
| No. of depressive episodes between child age 2 and 9 | 1.25 | 1.70 | 0 | 6 |
| Had at least one episode of depression | 0.48 |  | 0 | 1 |
| Recent depression | 0.24 |  | 0 | 1 |
| Mean EPDS | 5.60 | 3.80 | 0 | 24.33 |
| *Estimation Sample: Test-scores (age 14)* |  |  |  |  |
| No. of depressive episodes between child age 0 and 5 | 1.08 | 1.46 | 0 | 5 |
| No. of depressive episodes between child age 5 and 9 | 0.43 | 0.70 | 0 | 2 |
| No. of depressive episodes between child age 2 and 9 | 1.25 | 1.71 | 0 | 6 |
| Had at least one episode of depression | 0.48 |  | 0 | 1 |
| Recent depression | 0.23 |  | 0 | 1 |
| Mean EPDS | 5.62 | 3.84 | 0 | 24 |
| *Estimation Sample: Test-scores (age 16)* |  |  |  |  |
| No. of depressive episodes between child age 0 and 5 | 1.06 | 1.44 | 0 | 5 |
| No. of depressive episodes between child age 5 and 9 | 0.44 | 0.70 | 0 | 2 |
| No. of depressive episodes between child age 2 and 9 | 1.25 | 1.70 | 0 | 6 |
| Had at least one episode of depression | 0.48 |  | 0 | 1 |
| Recent depression | 0.24 |  | 0 | 1 |
| Mean EPDS | 5.56 | 3.76 | 0 | 24 |
| *Estimation Sample: Total SDQ (age 11)* |  |  |  |  |
| No. of depressive episodes between child age 0 and 5 | 1.03 | 1.44 | 0 | 5 |
| No. of depressive episodes between child age 5 and 9 | 0.43 | 0.70 | 0 | 2 |
| No. of depressive episodes between child age 2 and 9 | 1.21 | 1.70 | 0 | 6 |
| Had at least one episode of depression | 0.47 |  | 0 | 1 |
| Recent depression | 0.23 |  | 0 | 1 |
| Mean EPDS | 5.47 | 3.72 | 0 | 24 |
| *Estimation Sample: Total SDQ (age 13)* |  |  |  |  |
| No. of depressive episodes between child age 0 and 5 | 0.97 | 1.40 | 0 | 5 |
| No. of depressive episodes between child age 5 and 9 | 0.40 | 0.68 | 0 | 2 |
| No. of depressive episodes between child age 2 and 9 | 1.14 | 1.66 | 0 | 6 |
| Had at least one episode of depression | 0.45 |  | 0 | 1 |
| Recent depression | 0.22 |  | 0 | 1 |
| Mean EPDS | 5.30 | 3.60 | 0 | 24 |
| *Estimation Sample: Total SDQ (age 16)* |  |  |  |  |
| No. of depressive episodes between child age 0 and 5 | 0.95 | 1.38 | 0 | 5 |
| No. of depressive episodes between child age 5 and 9 | 0.39 | 0.66 | 0 | 2 |
| No. of depressive episodes between child age 2 and 9 | 1.10 | 1.62 | 0 | 6 |
| Had at least one episode of depression | 0.21 |  | 0 | 1 |
| Recent depression | 0.44 |  | 0 | 1 |
| Mean EPDS | 5.27 | 3.54 | 0 | 24 |

Note: These figures refer to the estimation samples used in Table A10.

**Table A10:** Alternative Measures of Maternal Depression and Child Human Capital: 2SLS results

|  | Test-scores |  |  | Total SDQ |  |  |
| --- | --- | --- | --- | --- | --- | --- |
|  | Age 11<br>(1) | Age 14<br>(2) | Age 16<br>(3) | Age 11<br>(4) | Age 13<br>(5) | Age 16<br>(6) |
| **Panel A (Main estimates from Table 2)** |  |  |  |  |  |  |
| No. of depressive episodes<br>between child ages 0 and 9 | -0.227*<br>(0.118) | -0.179*<br>(0.093) | -0.284***<br>(0.102) | -0.394***<br>(0.125) | -0.416***<br>(0.132) | -0.498**<br>(0.211) |
| F-statistics | 14.8 | 21.3 | 21.8 | 22.1 | 22.2 | 10.3 |
| Observations | 2601 | 2036 | 2828 | 2993 | 2585 | 2076 |
| **Panel B** |  |  |  |  |  |  |
| No. of depressive episodes<br>up to child's 5<sup>th</sup> birthday | -0.273**<br>(0.139) | -0.223*<br>(0.115) | -0.364***<br>(0.130) | -0.512***<br>(0.162) | -0.579***<br>(0.187) | -0.597**<br>(0.244) |
| F-statistics | 18.2 | 24.7 | 23.7 | 23.5 | 20.5 | 12.5 |
| Observations | 2601 | 2036 | 2828 | 2993 | 2585 | 2076 |
| **Panel C** |  |  |  |  |  |  |
| No. of depressive episodes<br>between child ages 5 and 9 | -0.681*<br>(0.409) | -0.545*<br>(0.320) | -0.777**<br>(0.328) | -1.019***<br>(0.375) | -0.969***<br>(0.335) | -1.750<br>(1.112) |
| F-statistics | 6.0 | 8.4 | 10.4 | 12.4 | 15.6 | 3.0 |
| Observations | 2601 | 2036 | 2828 | 2993 | 2585 | 2076 |
| **Panel D** |  |  |  |  |  |  |
| No. of depressive episodes<br>between child ages 2 and 9 | -0.282*<br>(0.148) | -0.218*<br>(0.114) | -0.345***<br>(0.126) | -0.468***<br>(0.151) | -0.475***<br>(0.152) | -0.572**<br>(0.245) |
| F-statistics | 12.6 | 18.8 | 19.1 | 20.4 | 21.9 | 10.0 |
| Observations | 2601 | 2036 | 2828 | 2993 | 2585 | 2076 |
| **Panel E** |  |  |  |  |  |  |
| Had at least one episode<br>of depression (dummy) | -0.948*<br>(0.502) | -0.843*<br>(0.457) | -1.234***<br>(0.463) | -1.555***<br>(0.502) | -1.631***<br>(0.532) | -1.520**<br>(0.610) |
| F-statistics | 12.6 | 14.6 | 17.0 | 20.9 | 20.2 | 14.8 |
| Observations | 2601 | 2036 | 2828 | 2993 | 2585 | 2076 |
| **Panel F** |  |  |  |  |  |  |
| Recent depression (dummy) | -3.218<br>(2.759) | -1.972<br>(1.339) | -2.565**<br>(1.283) | -3.162**<br>(1.358) | -2.826**<br>(1.107) | -7.374<br>(7.789) |
| F-statistics | 1.6 | 3.8 | 5.5 | 7.1 | 9.9 | 0.9 |
| Observations | 2601 | 2036 | 2828 | 2993 | 2585 | 2076 |
| **Panel G** |  |  |  |  |  |  |
| Average EPDS | -0.889<br>(0.568) | -0.583*<br>(0.336) | -1.038**<br>(0.485) | -1.220***<br>(0.443) | -0.961***<br>(0.315) | -2.048<br>(1.394) |
| F-statistics | 3.7 | 7.1 | 6.4 | 9.1 | 15.2 | 2.2 |
| Observations | 2559 | 1998 | 2783 | 2947 | 2548 | 2053 |

Notes: Standard errors appear in parentheses. All dependent variables are standardised. All non-binary measures of maternal depression are standardised. The controls are: the child's gender, birth year, birth order, number of siblings, the age of the mother at the birth of the child; dummies for the mother having at least an A-level, being employed, having a partner, having a partner with at least an A-level, having an employed partner, dummies for banded household income and ancestry-informative principal components. All of the regressions with test-scores as the dependent variables include school and school-year fixed effects. The difference in observations in the sixth panel is due to missing values for the EPDS (Edinburgh Postnatal Depression Score) when the cohort member was eight years old. The reported F-statistics are those for the Cragg-Donald Wald weak-identification test. \*\*\* p<0.01, \*\* p<0.05, \* p<0.1.

**Table A11:**
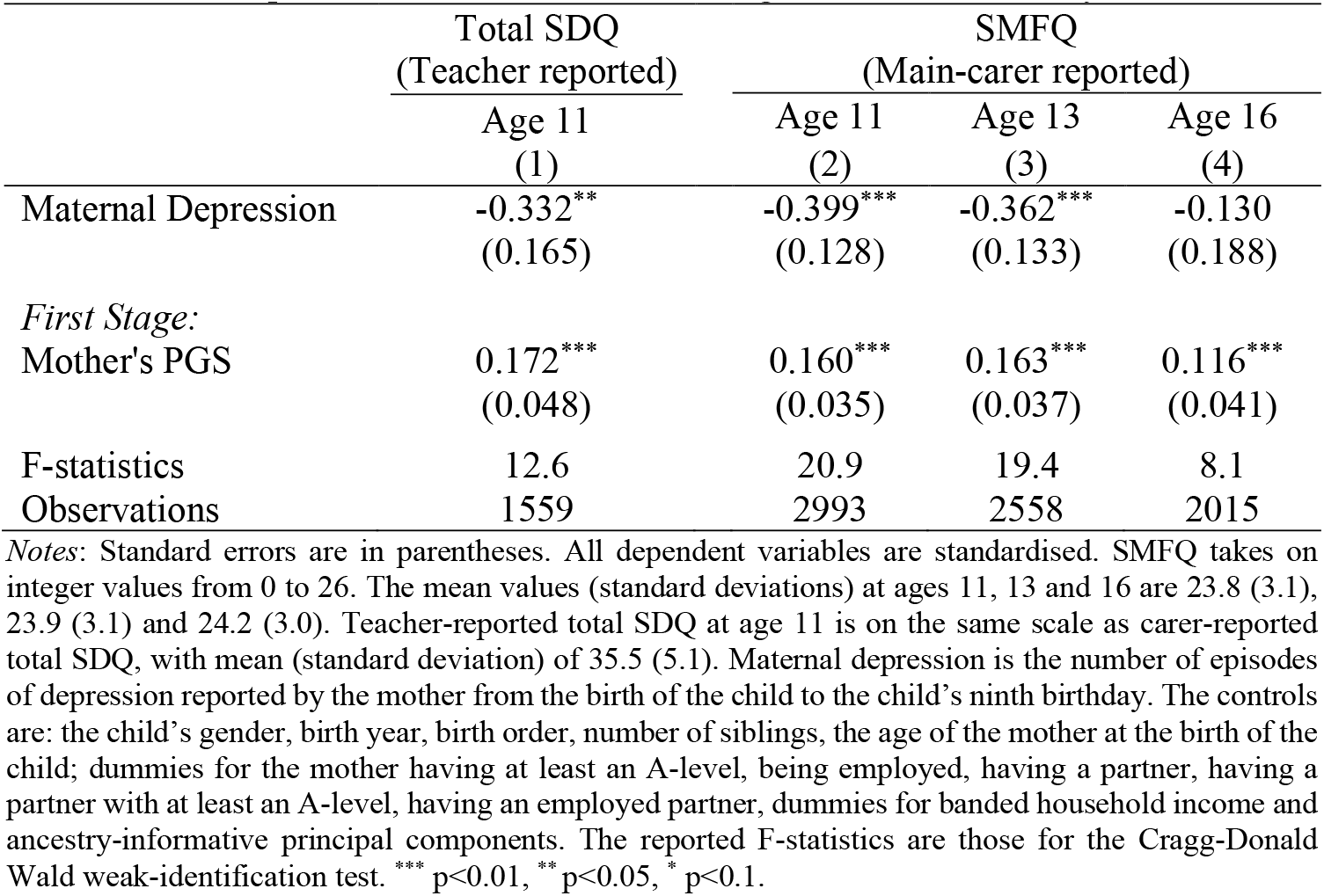
Maternal Depression and Children’s Non-Cognitive Skills: Validity Tests - 2SLS Results

**Table A12:** Short Moods and Feelings Questionnaire (SMFQ) *These questions are about how your teenager may have been feeling or acting recently*. *For each question, please say how much you think he/she has felt or acted this way in the past two weeks*.

| Your teenager: | Not True | Somewhat True | True |
| --- | --- | --- | --- |
| 1. Felt miserable or unhappy | 2 | 1 | 0 |
| 2. Didn't enjoy anything at all | 2 | 1 | 0 |
| 3. Felt so tired that he/she just sat around and did nothing | 2 | 1 | 0 |
| 4. Was very restless | 2 | 1 | 0 |
| 5. Felt he/she was no good anymore | 2 | 1 | 0 |
| 6. Cried a lot | 2 | 1 | 0 |
| 7. Found it hard to think properly or concentrate | 2 | 1 | 0 |
| 8. Hated himself/herself | 2 | 1 | 0 |
| 9. Was a bad person | 2 | 1 | 0 |
| 10. Felt lonely | 2 | 1 | 0 |
| 11. Thought nobody really loved him/her | 2 | 1 | 0 |
| 12. Thought he/she could never be as good as other kids | 2 | 1 | 0 |
| 13. Felt he/she did everything wrong | 2 | 1 | 0 |

## Appendix B: Polygenic scores

Genetic variants, such as single-nucleotide polymorphisms (SNPs), are locations in the human DNA at which a certain degree of variation is observed across individuals in a population. There are only two possible nucleotide variations for each SNP, which are called alleles. Some specific alleles, the “effect alleles”, are associated with particular diseases or traits (due to evolutionary mechanisms of natural selection, these are typically the alleles appearing less frequently in the population, i.e. the “minor alleles”). The number of effect alleles that an individual possesses for a given SNP (the so-called “allelic dosage”) can either be 0 (no effect allele), 1 (only one effect allele), or 2 (both alleles are the effect allele).

Polygenic scores (PGS) are weighted sums or averages of individual allelic dosages for a given set of SNPs. Both the weights and the set of relevant SNPs are obtained from the publicly-available summary statistics of an existing GWAS. These are typically tables or text files providing a list of SNPs that are robustly associated with a trait, accompanied by a range of characteristics (e.g. the effect size of the SNP-trait association, the p-value of such association, and the effect allele). For a given individual *j* in a prediction sample (independent of the training sample used in the GWAS), the default formula for the calculation of her PGS in the command line program PLINK 1.9 (www.cog-genomics.org/plink/) is:

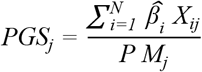

where 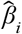 is the estimated effect size of SNP *i* on the trait of interest (obtained from a GWAS), *X*_*ij*_ the number of effect alleles observed in individual *j* for SNP *i, P* the ploidy of the sample (i.e. the number of sets of chromosomes in a cell, which is generally two for humans), *N* the total number of SNPs included in the PGS, and *M*_*j*_ the number of non-missing SNPs observed in individual *j*. If individual *j* has a missing genotype for SNP *i*, then the population minor-allele frequency multiplied by the ploidy (*MAF*_*i*_ *× P*) is used instead of *X*_*ij*_.

The allelic dosages of SNPs that are close to each other on a DNA strand tend to be correlated due to linkage disequilibrium (LD), i.e. *Cov (X*_*ij*_, *X*_*sj*_ ) ≠ *0* for *i* and *s* that are close enough. As each 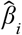 coming from the GWAS results is separately estimated from a linear regression of the trait of interest on SNP *i* in the training sample, some of the 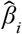 ‘s will be biased. Due to the overweighting of SNPs in long LD blocks, the resulting PGS will also be biased, and will have worse predictive accuracy. There are several ways to account for LD when interpreting the results of a GWAS: some, such as pruning or clumping,^20^ are based on recursive algorithms where only approximately-independent SNPs are retained for PGS construction; others, such as LDpred (Vilhjálmsson *et al*., 2015), use more complex machine-learning algorithms and Bayesian inference to obtain corrected effect-size estimates 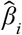 that take LD into account. It is increasingly common for GWAS authors to identify a “clean” set of approximately-independent SNPs (either via pruning or clumping). This is the case for the depression meta-analysis in Turley *et al*. (2018) that we use to calculate our instrumental variable. The single-trait meta-analysis expands on the SNPs for depressive symptoms already identified in Okbay *et al*. (2016), using a larger sample of 465,337 individuals from UK Biobank, 23andMe, and the Resource for Genetic Epidemiology Research on Adult Health and Aging (GERA). In particular, using clumping (for more details see the Online Methods of Turley *et al*., 2018), the single-trait depression GWAS of Turley and co-authors identified 88 SNPs for depression significant at least at a 10^−6^ p-value threshold, of which 68 were available in our ALSPAC genotyped data and used for the construction of the PGS for maternal depression.^21^ Out of those 88 SNPs, 32 have a p-value lower than the genome-wide significant threshold 5×10^−8^ and were used in robustness checks to test the sensitivity of our PGS to the number of SNPs included in its computation (available upon request).

Other PGSs used for the production of Table 3 were derived from the GWASs of Demange *et al*. (2021) and Middeldorp *et al*. (2016). The former, which captures the cognitive aspects of educational attainment, was used to calculate the PGS for cognitive skills. We clumped the GWAS summary statistics using p-value thresholds of, respectively, 5×10^−8^ for the lead SNPs and 10^−6^ for the SNPs in the clumps. Clumps were defined based on windows of 1000 kb from the lead SNPs and squared pairwise correlations of at least 0.1 (LD patterns were inferred from the sequenced genotypes of 379 individuals of European descent from Phase 1 of the 1000 Genomes Project). Only 179 out of the 225 lead SNPs were found in ALSPAC children and used for the calculation of the child’s PGS for cognitive skills.

For non-cognitive skills, we first applied the same clumping procedure to the SNPs that were significantly associated with the non-cognitive aspects of educational attainment from the GWAS-by-subtraction in Demange *et al*. (2021). However, the resulting PGS is not predictive of any of our measures of non-cognitive development in ALSPAC. This could reflect that the measure used in Demange *et al*. (2021) to determine non-cognitive skills (i.e. the portion of educational attainment that is not explained by cognitive skills) is very different from our measures of non-cognitive skills (the SDQ and SMFQ). Furthermore, with respect to cognitive ability, non-cognitive skills encompass a broader and harder-to-define variety of traits, which are also more likely to change over the life course. Consequently, the weights derived from GWASs in adult populations may not provide an appropriate summary of the importance of these genetic variants in the prediction of child non-cognitive outcomes. As such, we considered the GWASs of non-cognitive skills in populations of children and/or adolescents. The GWASs from the Early Genetics and Lifecourse Epidemiology (EAGLE) consortium are to our knowledge the only appropriate ones, as they use cohorts of children and adolescents to analyse internalising problems (Benke *et al*., 2014), ADHD, (Middeldorp *et al*., 2016) and aggressive behaviour (Pappa *et al*., 2016).^22^ As the sample sizes here are much smaller than those in adult populations GWASs, the SNP-trait associations are less-precisely estimated. We thus use less-stringent p-value thresholds to select a subset of approximately-independent SNPs via clumping: leading SNPs should have p-values of 5×10^−5^ at most, and only SNPs with a p-value lower than 0.001 can form the clumps. Other than that, the clumping procedure is the same as that described above for the depression PGS. After clumping, the PGS from the eight lead SNPs from the ADHD GWAS of Middeldorp *et al*. (2016) proved to be the most predictive of total SDQ across ages 11, 13 and 16. The PGSs from the clumped SNPs of the other two GWASs (Benke *et al*., 2014, and Pappa *et al*., 2016), although less-robustly associated with our measures of non-cognitive skills, were used for sensitivity analysis in Table 3 (results available upon request).

Last, our results throughout the paper do not depend on either clumping or the clumping p-value thresholds. The results remain qualitatively similar when using PGSs based on all SNPs above certain p-value thresholds, regardless of LD concerns.^23^

## Footnotes

1 The study website contains details of all the data that is available through a fully searchable data dictionary and variable search tool: http://www.bristol.ac.uk/alspac/researchers/our-data/.

2 At the end of Key Stages 2 (age 11) and 3 (age 14), children’s progress in Mathematics, Science and English is assessed using National Curriculum tests. While these tests do not produce exit certificates, the national exams taken at the end of Key Stage 4 (age 16), the General Certificate of Secondary Education (GCSE), do produce qualification certificates. Students in the UK typically take at least 5 GCSEs (one per subject), with Mathematics, Science and English being compulsory.

3 The Law of Independent Assortment does however come with a caveat: genes that are close to each other on a chromosome strand have a higher chance of being transmitted together. This leads to what is known as *linkage disequilibrium*: in a given population, alleles for different genes have higher association rates than those that would be expected from random matching.

4 Note that if parents were to match to each other independently of the trait that a given genotype regulates, we would not even need to condition on parental genotypes for the genotype of the child to be a random draw from the population genetic pool.

5 This stratification reflects drifts in allelic frequencies within the population of interest. A popular solution, which is particularly well-suited in contexts of considerable geographical and ethnic diversity, is controlling for principal components derived from genotyped data. These account for systematic associations between the alleles in subsets of a given population that are produced, among other things, by within-group assortative-matching patterns.

6 While the grandparents’ non-transmitted alleles can be unobserved confounders of the *D*_*M*_ - *Y*_*C*_ association (through genetic nurture, as they likely affect the way in which the grandparents bring up the mother), they do not play a role in the independence assumption of the effect of 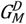 on *Y_C_*, as they cannot appear in the mother’s PGS for depression (Mendel’s Law of Segregation).

7 Note that controlling for 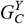 is what DiPrete *et al*. (2018) refer to as Unconditional Genetic Instrumental Variables (GIV-U), that is simply controlling for all the genetic variants associated with 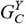. In particular, they show that GIV-U regression provides a reasonable lower bound for the true effect of *D*_*M*_ on *Y*_*C*_ under several violations of the IV assumptions (either through moderate pleiotropy or other genetic confounds).

8 An A-level (Advanced Level) qualification is a subject-based school-leaving certificate that is typically obtained at the end of Upper-Secondary School at around age 18.

9 The mother’s traits that are measured at child age eight (household income and the number of children in the household) are missing in roughly 9% of the cases in our different estimation samples. Where the respondents have missing information, we create a variable-specific dummy to flag this missing information (the Missing Indicator method) and replace the missing value by the sample mean. We in addition drop the 32 cases with multiple births for the mother over the 18-month initial survey period (although our results are robust to including these observations).

10 Technical details about the NPD cleaning process and the collection of the KS3 average grades we use here can respectively be found at http://www.bristol.ac.uk/media-library/sites/cmpo/migrated/documents/ks5userguide2011.pdf and https://find-npd-data.education.gov.uk/en/data_elements/11e50a8a-78d6-425c-871d-9d9fd3330dd9.

11 The reduced-form estimates for the PGS for depression range from -0.036 to -0.051 SD for cognitive skills and from -0.071 to -0.063 SD for non-cognitive skills, with significance levels identical to those in our baseline 2SLS estimates. While reduced-form estimates rely on weaker assumptions, they come at a cost in terms of interpretation, as they do not identify a mediating trait in the maternal genes - child outcome relationship. Under the assumptions described in Section 3.2, our 2SLS estimates reveal that the causal effect of the PGS for depression of the mother on the human child capital of children is only mediated by maternal depression.

12 We cannot make strong statements about whether the effect of maternal depression differs by gender, birth-order, maternal education and household-income band, as the smaller samples produce F-statistics that are mainly too low for robust inference.

13 Note that neither maternal depression nor the instrument predict retrospective attrition for cognitive skills in the top panel of Table 2. As information on Key Stages 2 and 3 (child ages 11 and 14, respectively) test-scores are obtained retrospectively, attrition here is the probability of being in the age-16 sample for cognitive skills and being absent from, respectively, the analogous age-11 and age-14 samples. The results on attrition in the cognitive and non-cognitive samples are available upon request.

14 It might be thought then that we would be better-off restricting our analysis only to episodes of maternal depression that are followed by a medical consultation. When doing so, we find coefficients that are on average twice as large as the baseline 2SLS estimates from Table 2 (all significant at least at the 10% level). However, the F-statistics for episodes of maternal depression followed by a medical visit take values that are systematically lower than those in Table 2. Using only depressive episodes followed by a medical visit comes at a greater risk of weak-instrument issues.

15 These are the following: rs10514301, rs10789340, rs10045971, rs11876620, rs12958048, and rs174548.

16 Note that the discovery sample of Middeldorp *et al*. (2016) includes the ALSPAC cohort. We also used alternative summary statistics from other GWAS (Benke *et al*., 2014; Pappa *et al*., 2016; Demange *et al*., 2021) to calculate alternative polygenic scores for non-cognitive skills, but none of these significantly correlates with total SDQ other than that from Middeldorp *et al*. (2016). These results are available upon request.

17 Another way of ruling out confounding genetic-inheritance effects is to recalculate the PGS for maternal depression excluding the genetic variants that are also associated with children’s cognitive and non-cognitive skills (either directly or through LD patterns). Out of the 68 top variants for maternal depression genotyped in ALSPAC, we find that none coincides with top variants for cognitive skills, while fourteen others are in LD with at least one cognitive top variant. In contrast, we find no overlap with the eight main genetic variants for non-cognitive outcomes (as measured by ADHD). The results, available upon request, remain qualitatively unchanged.

18 For the sake of transparency, the dashed grey lines in Figure A2 show the 90% confidence intervals when following the ‘local-to-zero’ approach described in van Kippersluis and Rietveld (2018), where λ is assumed to follow a Normal distribution and where there is no subsample for which the first-stage is zero. When we do so half of our baseline estimates, i.e. λ=0, are no longer significantly different from zero at the 10% level. Note that van Kippersluis and Rietveld (2018) apply this method to an estimation sample with over 100,000 observations. With roughly 3,000 observations at best, our estimation samples may well be too small to provide sufficient precision here.

19 Total SDQ can be split into two finer subscales: internalising SDQ (emotional health: the sum of ‘peer problems’ and ‘emotional problems’) and externalising SDQ (behavioural issues: the sum of ‘hyperactivity/inattention’ and ‘conduct problems’). Maternal depression produces worse outcomes for both internalising and externalising SDQ. These results are available upon request.

20 Pruning takes the available SNPs in the prediction sample as the starting point. For each SNP in a defined window, a pruning algorithm generally calculates the Variance Inflation Factor (VIF) or the squared pairwise correlation between each pair of SNPs, and removes the pair if the LD is greater than a certain threshold (e.g. 0.5). The procedure is then repeated shifting the window a certain number of SNPs forward. Clumping, on the other hand, uses the GWAS summary results as the starting point. This procedure starts by taking the SNP whose association with the trait of interest has the smallest p-value (the “lead” SNP) and constructing a symmetric window around it; SNPs in the window that have a squared pairwise correlation with the lead SNP above a certain cut-off are assigned to the lead SNP’s clump. The algorithm continues by taking the next-most significant SNP that is not yet assigned to a clump and repeating the above procedure until there are no more significant SNPs (based on user-defined significance thresholds; for large GWASs, the genome-wide significance p-value threshold of 5×10^−8^ is often used for lead SNPs). The clumped set of SNPs is the list of all lead SNPs.

21 The full list of SNPs is available in the supplementary material of Turley *et al*. (2018). Those we could not use, as they were not available in the genotyped data of ALSPAC participants, are the following: rs1806153, rs3806843, rs4799936, rs9291059, rs9813064, rs10172121, rs10965565, rs113092725, rs11643097, rs11663393, rs12501627, rs12515229, rs1520081, rs189383553, rs192796028, rs28383313, rs28567442, rs413130, rs7126679, rs9663959.

22 Note that the training samples here include children from the ALSPAC cohort, thus violating the standard non-overlapping condition between the training and prediction sample (as we simply use the derived PGSs as controls in Table 3, however, we do not believe that this constitutes a major problem in our context).

23 In detail, we used p-value thresholds of either 5×10^−8^ or 10^−6^ for the PGSs for depression and cognitive skills, and thresholds of 5×10^−5^ and 0.001 for all the PGSs for non-cognitive skills.

## Notes

### Competing Interest Statement

The authors have declared no competing interest.

### Funding Statement

The UK Medical Research Council and the Wellcome Trust (Grant ref: 217065/Z/19/Z) and the University of Bristol provide core support for ALSPAC. A comprehensive list of Grants funding is available on the ALSPAC website (http://www.bristol.ac.uk/alspac/external/documents/grant-acknowledgements.pdf). Genotype data were generated by Sample Logistics and Genotyping Facilities at Wellcome Sanger Institute and LabCorp (Laboratory Corporation of America) using support from 23andMe. Financial support from the Fonds National de la Recherche Luxembourg (Grants C19/SC/13650569 and 10949242) is gratefully acknowledged. Andrew Clark acknowledges financial support from the EUR grant ANR-17-EURE-0001.

### Author Declarations

Ethics approval for the study was obtained from the ALSPAC Ethics and Law Committee and the Local Research Ethics Committees. Consent for biological samples has been collected in accordance with the Human Tissue Act. Informed consent for the use of data collected via questionnaires and clinics was obtained from participants following the recommendations of the ALSPAC Ethics and Law Committee at the time.

## References

Angrist, J. D., and Pischke, J. S. (2008). Mostly Harmless Econometrics: An Empiricist’s Companion. Princeton University Press.

Banerjee, S., Chatterji, P., and Lahiri, K. (2017). “Effects of psychiatric disorders on labor market outcomes: A latent variable approach using multiple clinical indicators.” Health Economics, 26, 184–205.

Baranov, V., Bhalotra, S., Biroli, P., and Maselko, J. (2020). “Maternal depression, women’s empowerment, and parental investment: Evidence from a randomized controlled trial.” American Economic Review, 110, 824–59.

Benke, K. S., Nivard, M. G., Velders, F. P., Walters, R. K., Pappa, I., Scheet, P. A., … and Verhulst, F. C. (2014). “A genome-wide association meta-analysis of preschool internalizing problems.” Journal of the American Academy of Child and Adolescent Psychiatry, 53, 667–676.

Boef, A. G., Dekkers, O. M., and Le Cessie, S. (2015). “Mendelian randomization studies: a review of the approaches used and the quality of reporting.” International Journal of Epidemiology, 44, 496–511.

Boyd, A., Macleod, J., Henderson, J., Molloy, L., Ring, S., Golding, J., and Ness, A. (2013). “Cohort profile: The “Children of the 90s”-The index offspring of the Avon longitudinal study of parents and children.” International Journal of Epidemiology, 42, 111–127.

Boyle, E. A., Li, Y. I., and Pritchard, J. K. (2017). “An expanded view of complex traits: from polygenic to omnigenic.” Cell, 169, 1177–1186.

Briole, S., Le Forner, H., and Lepinteur, A. (2020). “Children’s socio-emotional skills: Is there a quantity–quality trade-off?” Labour Economics, 64, https://doi.org/10.1016/j.labeco.2020.101811.

Bubonya, M., Cobb-Clark, D. A., and Wooden, M. (2017). “Mental health and productivity at work: Does what you do matter?” Labour Economics, 46, 150–165.

Clark, A.E., D’Ambrosio, C., and Barazzetta, M. (2021). “Childhood circumstances and young adult outcomes: The role of mothers’ financial problems”. Health Economics, 30, 342–357.

Clark, A.E., Flèche, S., Layard, R., Powdthavee, N., and Ward, G. (2018). The Origins of Happiness: The Science of Well-being over the Life Course. Princeton University Press.

Clark, A.E., and Lepinteur, A. (2019). “The causes and consequences of early-adult unemployment: Evidence from cohort data.” Journal of Economic Behavior & Organization, 166, 107–124.

Clark, D. M. (2018). “Realizing the mass public benefit of evidence-based psychological therapies: the IAPT program.” Annual Review of Clinical Psychology, 14, 159–183.

Conley, T. G., Hansen, C. B., and Rossi, P. E. (2012). “Plausibly exogenous.” Review of Economics and Statistics, 94, 260–272.

Cunha, F., and Heckman, J. J. (2008). “Formulating, identifying and estimating the technology of cognitive and noncognitive skill formation.” Journal of Human Resources, 43, 738–782.

Dahlen, H. M. (2016). “The impact of maternal depression on child academic and socioemotional outcomes.” Economics of Education Review, 52, 77–90.

Davey Smith, G., and Hemani, G. (2014). “Mendelian randomization: Genetic anchors for causal inference in epidemiological studies.” Human Molecular Genetics, 23, 89–98.

Davies, N. M., von Hinke Kessler Scholder, S., Farbmacher, H., Burgess, S., Windmeijer, F., and Smith, G. D. (2015). “The many weak instruments problem and Mendelian randomization.” Statistics in Medicine, 34, 454–468.

Del Bono, E., Kinsler, J., and Pavan, R. (2020). Skill Formation and the Trouble with Child Non-Cognitive Skill Measures. IZA Discussion Paper No. 13713.

Demange, P. A., Malanchini, M., Mallard, T. T., Biroli, P., Cox, S. R., Grotzinger, A. D., … and Corcoran, D. (2021). “Investigating the genetic architecture of non-cognitive skills using GWAS-by-subtraction.” Nature Genetics, 53, 35–44.

DiPrete, T. A., Burik, C. A., and Koellinger, P. D. (2018). “Genetic instrumental variable regression: Explaining socioeconomic and health outcomes in nonexperimental data.” Proceedings of the National Academy of Sciences, 115, 4970–4979.

Flèche, S. (2017). Teacher Quality, Test-scores and Non-Cognitive Skills: Evidence from Primary School Teachers in the UK. CEP Discussion Paper No. 1472.

Fletcher, J. (2013). “Adolescent depression and adult labor market outcomes.” Southern Economic Journal, 80, 26–49.

Fraser, A., Macdonald-Wallis, C., Tilling, K., Boyd, A., Golding, J., Davey Smith, G., … and Ring, S. (2013). “Cohort profile: The Avon Longitudinal Study of Parents and Children: ALSPAC mothers cohort”. International Journal of Epidemiology, 42, 97–110.

Goodman, R. (1997). “The Strengths and Difficulties Questionnaire: A research note.” Journal of Child Psychology and Psychiatry, 38, 581–586.

Goodman, A., Lamping, D. L., and Ploubidis, G. B. (2010). “When to use broader internalising and externalising subscales instead of the hypothesised five subscales on the Strengths and Difficulties Questionnaire (SDQ): Data from British parents, teachers and children.” Journal of Abnormal Child Psychology, 38, 1179–1191.

Goodman, S. H., Rouse, M. H., Connell, A. M., Broth, M. R., Hall, C. M., and Heyward, D. (2011). “Maternal depression and child psychopathology: A meta-analytic review.” Clinical Child and Family Psychology Review, 14, 1–27.

Gotlib, I., Goodman, S., and Humphreys, K. (2020). “Studying the Intergenerational Transmission of Risk for Depression: Current Status and Future Directions.” Current Directions in Psychological Science, 29, 174–179.

Gotlib, I. H., Lewinsohn, P. M., and Seeley, J. R. (1998). “Consequences of depression during adolescence: Marital status and marital functioning in early adulthood.” Journal of Abnormal Psychology, 107, 686–690.

Grogger, J., and Eide, E. (1995). “Changes in college skills and the rise in the college wage premium.” Journal of Human Resources, 30, 280–310.

Hakulinen, C., Elovainio, M., Arffman, M., Lumme, S., Pirkola, S., Keskimäki, I., … and Böckerman, P. (2019). “Mental disorders and long-term labour market outcomes: Nationwide cohort study of 2,055,720 individuals.” Acta Psychiatrica Scandinavica, 140, 371–381.

Hansell, N. K., Halford, G. S., Andrews, G., Shum, D. H., Harris, S. E., Davies, G., … and Medland, S. E. (2015). “Genetic basis of a cognitive complexity metric”. PloS One, 10, https://doi.org/10.1371/journal.pone.0123886.

Hanushek, E.A. and Kimko, D.D. (2000). “Schooling, labor-force quality and the growth of nations.” American Economic Review, 90, 1184–1208.

Heckman, J. J., Stixrud, J., and Urzua, S. (2006). “The effects of cognitive and noncognitive abilities on labor market outcomes and social behavior.” Journal of Labor Economics, 24, 411–482.

Heckman, J. J., Humphries, J. E., and Veramendi, G. (2018). “Returns to education: The causal effects of education on earnings, health, and smoking.” Journal of Political Economy, 126, 197–246.

Hemani, G., Bowden, J., and Davey Smith, G. (2018). “Evaluating the potential role of pleiotropy in Mendelian randomization studies.” Human Molecular Genetics, 27, 195–208.

Karlsson Linnér, R., Biroli, P., Kong, E., Meddens, S. F. W., Wedow, R., Fontana, M. A., … and Nivard, M. G. (2019). “Genome-wide association analyses of risk tolerance and risky behaviors in over 1 million individuals identify hundreds of loci and shared genetic influences.” Nature Genetics, 51, 245–257.

Kiernan, K. E., and Huerta, M. C. (2008). “Economic deprivation, maternal depression, parenting and children’s cognitive and emotional development in early childhood.” British Journal of Sociology, 59, 783–806.

Koellinger, P. D., and De Vlaming, R. (2019). “Mendelian randomization: the challenge of unobserved environmental confounds.” International Journal of Epidemiology, 48, 665–671.

Kong, A., Thorleifsson, G., Frigge, M. L., Vilhjalmsson, B. J., Young, A. I., Thorgeirsson, T. E., … and Gudbjartsson, D. F. (2018). “The nature of nurture: effects of parental genotypes.” Science, 359, 424–428.

Lawlor, D., Richmond, R., Warrington, N., McMahon, G., Smith, G. D., Bowden, J., and Evans, D. M. (2017). “Using Mendelian randomization to determine causal effects of maternal pregnancy (intrauterine) exposures on offspring outcomes: Sources of bias and methods for assessing them.” Wellcome Open Research, 2, https://doi.org/10.12688/wellcomeopenres.10567.1.

Lee, J. J., Wedow, R., Okbay, A., Kong, E., Maghzian, O., Zacher, M., … & Fontana, M. A. (2018). “Gene discovery and polygenic prediction from a genome-wide association study of educational attainment in 1.1 million individuals.” Nature Genetics, 50, 1112–1121.

Middeldorp, C. M., Hammerschlag, A. R., Ouwens, K. G., Groen-Blokhuis, M. M., Pourcain, B. S., Greven, C. U., … and Vilor-Tejedor, N. (2016). “A genome-wide association meta-analysis of attention-deficit/hyperactivity disorder symptoms in population-based pediatric cohorts.” Journal of the American Academy of Child and Adolescent Psychiatry, 55, 896–905.

Murnane, R.J., Singer, J.D., Willet, J.B., Kemple, J.J. and Olsen, R. (1991). Who Will Teach? Policies that Matter. Cambridge, MA: Harvard University Press.

Nybom, M. (2017). “The distribution of lifetime earnings returns to college.” Journal of Labor Economics, 35, 903–952.

O’Hara, M. W., and McCabe, J. E. (2013). “Postpartum depression: Current status and future directions.” Annual Review of Clinical Psychology, 9, 379–407.

Okbay, A., Baselmans, B. M., De Neve, J. E., Turley, P., Nivard, M. G., Fontana, M. A., … and Gratten, J. (2016). “Genetic variants associated with subjective well-being, depressive symptoms, and neuroticism identified through genome-wide analyses.” Nature Genetics, 48, 624–633.

Pappa, I., St Pourcain, B., Benke, K., Cavadino, A., Hakulinen, C., Nivard, M. G., … and Evans, D. M. (2016). “A genome-wide approach to children’s aggressive behavior: The EAGLE consortium.” American Journal of Medical Genetics Part B: Neuropsychiatric Genetics, 171, 562–572.

Perry, C. D. (2008). “Does treating maternal depression improve child health management? The case of pediatric asthma.” Journal of Health Economics, 27, 157–173.

Prince, M., Patel, V., Saxena, S., Maj, M., Maselko, J., Phillips, M.R., and Rahman, A. (2007). “No health without mental health.” Lancet, 370, 859–877.

Rajagopal, V. M., Ganna, A., Coleman, J. R., Allegrini, A. G., Voloudakis, G., Grove, J., … and Schork, A. (2020). Genome-wide association study of school grades identifies a genetic overlap between language ability, psychopathology and creativity. bioRxiv.

Smith, G. D., Lawlor, D. A., Harbord, R., Timpson, N., Day, I., and Ebrahim, S. (2007). “Clustered environments and randomized genes: A fundamental distinction between conventional and genetic epidemiology”. PLoS Medicine, 4, 1985–1992.

Stansfeld, S., Clark, C., Bebbington, P., King, M., Jenkins, R., and Hinchliffe, S. (2016). “Chapter 2: Common mental disorders.” In S. McManus, P. Bebbington, R. Jenkins, and T. Brugha (Eds.), Mental Health and Wellbeing in England: Adult Psychiatric Morbidity Survey 2014 (pp. 37–68). Leeds: NHS Digital.

Taylor, A. E., Jones, H. J., Sallis, H., Euesden, J., Stergiakouli, E., Davies, N. M., … & Tilling, K. (2018). “Exploring the association of genetic factors with participation in the Avon Longitudinal Study of Parents and Children.” International Journal of Epidemiology, 47, 1207–1216.

Turley, P., Walters, R. K., Maghzian, O., Okbay, A., Lee, J. J., Fontana, M. A., … and Magnusson, P. (2018). “Multi-trait analysis of genome-wide association summary statistics using MTAG.” Nature Genetics, 50, 229–237.

Van Kerm, P. (2003). “Adaptive kernel density estimation”. Stata Journal, 3, 148–156.

Van Kippersluis, H., and Rietveld, C. A. (2018). “Pleiotropy-robust Mendelian randomization.” International Journal of Epidemiology, 47, 1279–1288.

Von Hinke, S., Rice, N., and Tominey, E. (2019). Mental Health around Pregnancy and Child Development from Early Childhood to Adolescence. IZA Discussion Paper No. 12544.

Von Hinke, S., Smith, G. D., Lawlor, D. A., Propper, C., and Windmeijer, F. (2016). “Genetic markers as instrumental variables.” Journal of Health Economics, 45, 131–148.

Zhang, G., Bacelis, J., Lengyel, C., Teramo, K., Hallman, M., Helgeland, Ø., … and Jacobsson, B. (2015). “Assessing the causal relationship of maternal height on birth size and gestational age at birth: a mendelian randomization analysis.” PLoS Medicine, 12, 10.1371/journal.pmed.1001865.

Zimmerman F., and Katon, W. (2005). “Socioeconomic status, depression disparities, and financial strain: What lies behind the income-depression relationship?” Health Economics, 14, 1197–1215.

## References for Appendix B

Vilhjálmsson, B. J., Yang, J., Finucane, H. K., Gusev, A., Lindström, S., Ripke, S., ... & Hayeck, T. (2015). “Modeling linkage disequilibrium increases accuracy of polygenic risk scores.” American Journal of Human Genetics, 97, 576–592.

